# Quantifying Cognitive Reserve Through Structural-Functional Interactions: Neuroadaptive Biomarkers in Aging and Neurodegenerative Pathologies

**DOI:** 10.1101/2025.08.25.25334345

**Authors:** Yumeng Li, Xinyue Zhang, Xin Li, Zhanjun Zhang

## Abstract

**Background:** Cognitive reserve (CR) explains individual resilience to age-related cognitive decline, yet its neurobiological basis remains elusive. Current CR proxies lack direct mechanistic links, necessitating a system-level approach integrating brain structure-function interactions.

**Methods:** We developed a novel CR metric using structural MRI and resting-state fMRI from 1,280 older adults. A youth-derived structural-functional prediction model estimated maximal attainable brain function in elders. CR was quantified as the deviation between observed and predicted function. Cross-sectional and longitudinal analyses assessed CR’s spatial distribution, cognitive associations, and pathological relevance in MCI/AD cohorts.

**Results:** CR hubs localized to prefrontal, cingulate, and precuneus regions, organized within high-order networks. Higher CR predicted slower cognitive decline (r = −0.21, p < 0.001) and correlated with reduced Aβ deposition (r = −0.63, p < 0.001). CR demonstrated domain-specific associations with memory, attention, and processing speed. MCI exhibited broader CR reductions than AD, particularly in frontotemporal regions, likely reflecting stage-specific neuroplastic dynamics: early MCI retains partial compensatory capacity but inefficient CR utilization under mounting pathological stress, whereas advanced AD transitions to irreversible structural damage that disrupts CR’s adaptive “software” mechanisms.

**Conclusions:** This study establishes CR as a dynamic neuroprotective framework, bridging functional resilience and structural integrity. CR’s spatial specificity and inverse link to amyloid pathology highlight its potential as an early biomarker for resisting pathological aging.

## 1 Introduction

CR refers to the adaptive characteristics of cognitive processes, such as efficiency, capacity, and flexibility --that account for variability in cognitive function amid brain aging, pathology, or injury[2–4]. Its core lies in the dynamic adjustment of functional brain processes rather than deliberate activation. This reserve model enables individuals to cope with brain structural changes through compensatory mechanisms. The relevant neural mechanisms may precede pathology. Also, differences in cognitive adaptation among individuals significantly impact their capacity to combat brain aging or diseases [2, 5, 6].

Thus, accurately quantifying cognitive reserve (CR) is vital for probing individual differences in late - life development and uncovering mechanisms underlying cognitive decline. Current CR quantification methods include: 1. Proxy indicators of social behavior: Metrics such as education, occupational complexity, socioeconomic status, and leisure activities serve as indirect proxies for CR [7–11]. While these factors reflect experiences that contribute to CR development, they lack direct links to specific CR mechanisms and are limited in explaining CR’s neurobiological basis. 2. Residual-based quantification: This method builds a cognitive performance prediction model with demographic indicators and brain-based predictors [12]. The unexplained cognitive variance in the model is used as the quantitative CR index. Its validity depends largely on the proper selection of predictors and outcome variables in the model. Note that brain measures used for cognitive prediction may only partially capture the brain’s physiological and pathological features. Like proxy indicators of social behaviors, they risk including extra factors not fitting the reserve definition. Also, due to variable - set differences across studies, results are bound to be more variable [2, 13–15]. 3. Quantifying CR using functional imaging: Some studies indicate that brain functional network properties are crucial for capturing CR and may serve as its neurobiological basis [16, 17]. These network properties correlate with typical social-behavioral proxy factors of CR and can mediate the impact of brain changes on cognition [18–20]. Based on this, some experts suggest that identifying and verifying such brain networks could provide a more direct and accurate measure of an individual’s CR.

Current evidence suggests that functional neuroimaging represents the most promising approach for quantifying cognitive reserve (CR). However, relying solely on brain function to interpret CR remains limited, as its direct mechanistic expression in individuals originates from micro-scale neuronal processing mechanisms [21–23]. While this mechanism transcends the constraints of task-based fMRI, exhibiting greater generalizability and adaptability, large-scale direct detection of neuronal responses remains a formidable technical challenge. Consequently, integrating CR with molecular and cellular neurobiological mechanisms is highly complex. Hence, our research goal is to find a counterpart of CR from a system-wide perspective. This counterpart should bridge the gap between macro brain functional networks and micro neuronal mechanisms, offering a more comprehensive and precise framework for quantifying CR.

Brain structure, particularly gray matter surface structure, is crucial for shaping neuronal signal transmission [24–27]. These structural features determine the physical characteristics of neural signal propagation, giving rise to complex patterns of neuronal co-activation. These highly ordered and dynamic activation patterns are considered key neural mechanisms supporting perception, cognition, and various psychological functions[28]. In the human brain, structural and functional decline in specific regions is not random but shows significant spatiotemporal synchrony [29, 30]. That is, within similar time frames, the degradation of structural integrity and the reduction of functional efficiency in specific brain regions often occur together. Their interrelated evolution in particular brain areas jointly drives the overall trend of age-related cognitive changes, influencing cognitive performance and psychological states in old age.

The co-evolution of brain structure and function may indicate individual differences during aging. Within the cognitive reserve (CR) framework, brain reserve (BR), reflected in the quantity and quality of neurons, synapses, and neural pathways, a is neurobiological resource linked to CR and structural integrity [31, 32]. This microscopic neural infrastructure offers a stable resource base, helping individuals maintain cognitive function amid neurodegeneration or brain damage [2]. For instance, seniors with superior cognitive performance show high anatomical and physiological similarity to younger brains in structural integrity, neurotransmitter efficiency, and neural connectivity. Based on these features, cross-sectional studies can also help us understand individual differences in cognitive aging.

To more accurately quantify cognitive reserve (CR), we should build on functional brain imaging methods by including the interaction between brain structure and function. CR is about how individuals handle age - related or disease - linked brain changes. People with high CR can maintain decent cognitive function despite brain structure damage from aging or disease.

This study presents a new CR quantification method. Firstly, we established a “youthful” brain reference model using individuals aged 18-20y. The primary objective of establishing a youthful brain reference model is to benchmark optimal structure-function coupling during neurodevelopmental maturity, thereby isolating active CR mechanisms by quantifying deviations in older adults’ functional profiles from model predictions. Then, we used this model to predict the maximum brain function supported by the gray - matter cortex of older people. By comparing the actual and predicted brain function in seniors, we can identify CR and declining regions. This helps us understand how CR works in cognitive aging. This approach directly reflects neuroadaptive capacity to compensate for structural decline through functional reorganization, while excluding confounds from passive aging processes.

Our CR indicator, based on structure - function interaction, has shown the spatial brain - map features and group characteristics of CR values in seniors. Building on the interplay between brain structure and function, this study aims to establish a novel framework for quantifying cognitive reserve (CR) by leveraging a youth-referenced model to predict age-resilient functional capacity. We seek to (1) delineate spatially consistent CR networks underpinning cognitive reserve, (2) characterize age- and sex-specific reconfiguration of these networks across healthy aging trajectories, (3) validate their predictive utility for longitudinal cognitive decline and domain-specific performance, and (4) identify CR-driven neuroprotective signatures that distinguish pathological aging (MCI/AD) from normal decline while forecasting Aβ deposition patterns. By integrating structural constraints with functional adaptability, our approach provides a biologically grounded CR metric to unravel mechanisms of cognitive heterogeneity in aging and inform early intervention strategies.

## 2 Methods

### 2.1 Participants

Participants were from the BABRI community cohort [33], a community-based cohort of middle-aged and older adults started in 2008. The BABRI project was approved by the Ethics Committee of the State Key Laboratory of Cognitive Neuroscience and Learning (IRB_A_0010_2024001) and the Institutional Review Board of the Brain Imaging Center of Beijing Normal University. All participants gave written informed consent.

Inclusion criteria were: (1) aged ≥45 years; (2) ≥6 years of education; (3) no diagnosis of neurodegenerative (e.g., AD, Parkinson’s disease), neurological (e.g., severe cerebrovascular disease, brain tumor, trauma), or severe psychiatric disorders (e.g., major depression, bipolar disorder, schizophrenia); (4) no history of drug/alcohol abuse; (5) MMSE - Chinese version score ≥24 to exclude suspected dementia [34]; (6) physical and mental ability to complete neuropsychological tests and information collection; and (7) completed T1 - weighted MRI and resting - state fMRI scans with no contraindications and acceptable image quality.

Initially, 1,482 participants aged 50 - 90 were included. After excluding those who failed to meet the criteria and those with poor - quality images (incomplete scans, significant artifacts, fMRI maximum head motion > 3 mm/3°, or Mean FD > 0.5 mm), 1,280 participants remained (age: 66.45 ±7.26 years; 494 men, 786 women; education: 11.49 ±3.14 years). Among them, 323 had longitudinal cognitive assessment data (average follow-up: 3.19 ±1.34 years).

The participant group for studying disease-related cognitive reserve characteristics was separate from the above group. A total of 135 participants were recruited, including 76 NC, 30 MCI patients, and 29 AD patients. All participants completed florbetapir PET, structural MRI, and an extensive cognitive battery within a month. All participants met the following criteria: right-handed; native Chinese speakers; at least 50 years old; and no history of neurological or psychiatric disorders. The diagnostic criteria for MCI based on Petersen’s criteria included (1) the presence of subjective cognitive complaints (self-reported and/or by informants); (2) normal general cognition (a score higher than 23 on the Mini-mental State Examination [MMSE]); (3) intact daily function (a score of 0 in the Instrumental Activity of Daily Living [IADL] and ADL); and (4) objective cognitive impairment (neuropsychological tests scores in “Neuropsychological test” less than 1.5 standard deviations below the age- and education-adjusted norms of the Chinese elderly population) (Petersen 2010). AD was diagnosed according to the criteria of the National Institute of Neurological and Communicative Disorders and Stroke and the Alzheimer’s Disease and Related Disorders Association Dementia [35, 36] and further evaluated by brain CT or MRI. All participants gave written informed consent to our protocol, which was approved by the ethics committee of the State Key Laboratory of Cognitive Neuroscience and Learning, Beijing Normal University.

### 2.2 Behavioral and cognitive assessment

As described in our previous study, all participants underwent a battery of neuropsychological tests at baseline recruitment [33]. The assessment involved general cognitive ability and cognitive function across five domains including memory, language, attention, spatial processing, and executive function. General cognitive ability was tested using the Chinese version of the MMSE [34]. Memory was tested using the Auditory Verbal Learning Test (AVLT) [37]. Executive function was tested using the Stroop Color Word Test (SCWT) [38] and the Trail Making Test (TMT-B) [39]. Spatial processing was assessed using the Clock Drawing Test (CDT) [40]. Attention was evaluated using the Symbol Digit Modification Test (SDMT) [41]and the TMT_A test [39]. Language was tested using the Boston Naming Test (BNT) and the Verbal Fluency Test (VFT) [42].

### 2.3 MRI image acquisition and data processing

MRI data were acquired using a SIEMENS PRISMA 3T scanner at the Imaging Center for Brain Research at Beijing Normal University. Participants were in a supine position with their heads snugly fixed by straps and foam pads to minimize head movement. The T1-weighted structural images were acquired using 3D magnetization-prepared rapid gradient echo sequences: 192 sagittal slices, repetition time (TR)=2530 ms, echo time (TE)=2.27 ms, slice thickness=1 mm, flip angle (FOA)=7°, and field of view (FOV)=256 mm×256 mm.

The MATLAB2021b (https://ww2.mathworks.cn/) and SPM12 (https://www.fil.ion.ucl.ac.uk/spm/software/spm12/) toolboxes with default parameters were used to pre-process the structural images. The modulated GM images were smoothed with a Gaussian kernel of 8 mm full width at half maximum (FWHM). We utilized the mean GM map (threshold=0.2) of all the participants to obtain a group brain mask, as well as for subsequent analysis.

For all subjects, cortical reconstruction of T1-weighted images was performed using FreeSurfer version 5.3 (http://surf er.nmr.mgh.harvard.edu)[43]. This process involved registration to a template, intensity normalization, gray/white matter segmentation, and tessellation of gray/CSF and white/gray boundaries. Cortical surfaces were inflated and normalized via spherical registration, with cortical thickness defined as the shortest distance between the pial and white matter surfaces. The average regional cortical thickness was calculated without manual correction. The procedure included bias field correction, intensity normalization, and skull stripping using a watershed algorithm, followed by white matter segmentation, surface definition, and topology correction of the reconstructed surfaces. We used a triangular surface mesh representation of the midthickness human cortical surface. This representation, comprising 32,492 vertices in each hemisphere, was obtained from a down sampled, left–right symmetric version of the FreeSurfer’s fsaverage population-averaged template(https://github.com/ThomasYeoLab/CBIG/tree/master/data/templates/surface/ <u>fs_LR_164k</u>). Note that the template is independent of the data sample used in our analyses, thus avoiding concerns about circularity.

### 2.4 The preprocessing pipeline for functional neuroimaging data

Resting-state fMRI data were preprocessed using fMRIPrep (https://fmriprep.org/, V20.1.3), a Python-based automated tool integrating advanced neuroimaging packages like FSL, ANTs, FreeSurfer, and AFNI, designed for optimized fMRI preprocessing [44].

The procedure included head motion estimation, slice - timing correction, fMRI-to-T1w registration, fMRI-to-MNI152 standard - space normalization, confound estimation, and regression. Specifically, head motion parameters were estimated and saved for subsequent regression. Slice-timing correction adjusted for differences in slice acquisition times. The fMRI data were coregistered to the T1 - weighted structural image for precise alignment and then normalized to the MNI152 space for group - level analysis. Confounds such as head motion parameters, white matter, and CSF signals were estimated for regression. Post - preprocessing steps comprised linear drift removal, Gaussian smoothing (6 mm FWHM), and nonlinear band - pass filtering (0.01 - 0.1 Hz). Quality control involved visually checking T1w images and excluding subjects with excessive head motion (maximum head motion > 3 mm or 3°, or mean FD > 0.5 mm). All analyses were conducted at the voxel level to ensure accuracy and consistency.

### 2.5 PET processing

We used SPM12 to rigidly coregister the florbetapir PET scan to the structural MRI scan and spatially normalize it to a reference template. All PET images underwent smoothing with an 8 mm FWHM Gaussian filter. The standardized uptake value ratio (SUVR), a common semiquantitative method in antiamyloid drug trials, measures amyloid accumulation. Typically, the whole cerebellum or cerebellar gray matter serves as the reference region due to its minimal amyloid buildup [45–50]. Using the cerebellum as the reference region is based on the assumption that its amyloid accumulation is negligible. However, post-mortem studies have shown that a diffuse type of senile plaques exists in the cerebellum of some AD patients [46]; meanwhile, the longitudinal study found the subcortical white matter, such as corpus callosum was relatively unaffected by amyloid accumulation [51], and it has been shown to be a reliable reference brain region [50, 52]. The use of the cerebellum as the reference region in these cases where there is significant amyloid accumulation in the cerebellum is therefore inadequate. Thus, in the present study, we combined cerebellar GM and reliable reference brain region from longitudinal studies, that is, corpus callosum to generate average SUVR, from which the whole cerebellum was obtained from AAL, and the corpus callosum atlas came from the IIT Human Brain Atlas(https://www.nitrc.org/projects/iit)[52, 53].

### 2.6 Statistical analysis

#### Large Sample Dataset of Healthy Aging

##### 2.6.1 Cortical Reconstruction and Thickness Extraction

This study employed multimodal neuroimaging data processing to analyze the relationship between cortical thickness and functional reorganization. For structural data, we used FreeSurfer to reconstruct individual cortical thickness data and mapped it to the fsaverage space (163,842 vertices per hemisphere) via custom spherical registration, forming a vertex × participant matrix. For functional data, resting - state fMRI data was nonlinearly registered to fsaverage space, with preprocessing including temporal slice - timing correction, head motion correction (framewise displacement < 0.2 mm), and frequency - domain filtering (0.01 - 0.1 Hz), and then a three - dimensional functional matrix of vertex × time point × participant was built (Figure1.a).

##### 2.6.2 Measurement of CR Values

In the cortical functional reorganization analysis, machine learning-based predictive models were employed to establish associations between vertex-level cortical thickness and functional activity patterns. Specifically, vertex-level thickness values were utilized as input features for a decision tree regression algorithm. For each temporal time point, the model was trained using leave-one-out cross-validation (LOOCV) to ensure generalizability, with model reliability confirmed by achieving a prediction accuracy threshold of ≥60% when predicting functional activity from cortical thickness features in young participants.

Subsequently, the validated structural-functional prediction model derived from the young cohort was applied to the elderly cohort to estimate their maximally attainable functional capacity based on cortical structural integrity. Time-point-specific functional connectivity matrices were reconstructed by projecting the elderly participants’ cortical thickness data into the model. Utilizing the Schaefer2018 functional network parcellation template[54], time-series data for vertices corresponding to each network parcel were extracted. The Frobenius norm of the matrix difference between empirically observed and model-reconstructed functional connectivity matrices was computed to quantify global reconstruction error. Higher reconstruction errors (i.e., greater deviations from predicted function) indicated higher CR, reflecting diminished capacity to maintain functional performance despite structural aging (Figure1.b).

##### 2.6.3 SCCA Analysis

The current study used Sparse Canonical Correlation Analysis (SCCA) to explore the relationship between the spatial distribution of brain reorganization metrics (CR) and multi-domain cognitive function. Unlike traditional CCA, which struggles with high-dimensional data, SCCA introduces regularization to handle cases where feature dimensions exceed the sample size. It has been successfully applied in neuroscience research linking neuroimaging and behavioral data.

The analysis was performed in R using the PMA package. The model maximized the covariance between linear combinations of cortical thickness (u) and cognitive scores (v), while maintaining sparsity. This multivariate method effectively identifies key associations between brain reorganization and cognitive function.

To avoid overfitting and inflated covariance estimates, the study adopted a training/test split. Optimal sparsity parameters were determined via five-fold cross validation and grid search on the training set, with model performance validated on the independent test set. CR values and cognitive function canonical correlation coefficients were generated for the test set. Bonferroni correction was applied to control for multiple comparison false positives, with only features having |weights| > 0.1 reported. This design provides an unbiased estimate of CR-cognition associations.

##### 2.6.4 Covariate Control and Group Comparison

We controlled for the effects of covariates on the primary dependent variable, CR values, using multiple linear regression and extracted the residuals as latent variables adjusted for demographic factors. Specifically, we extracted age, sex, and years of education as covariates and centered them. For each brain region, we performed multiple linear regression with CR values as the dependent variable and age, sex, and education as independent variables. The residuals from this model were used for further analysis.

When comparing CR values between demographic groups (e.g., males vs. females), the covariates included only age and education. Similarly, when examining the effects of age and education on CR, the covariates were the remaining variables. Subsequently, we used independent-sample t-tests to compare the CR residuals between groups for the relevant brain regions (Figure1.c.①).

#### Cognitive Behavioral Longitudinal Dataset

##### 2.6.5 Calculating Rates of Cognitive Decline

Within the large sample of 1,280 participants, 323 had longitudinal cognitive - assessment data. For each individual, the rate of decline in each cognitive domain was calculated by subtracting baseline cognitive scores from follow - up scores and dividing by the time interval (in years) between baseline and follow - up.

##### 2.6.6 Correlations and Spatial Distribution of CR Values and Cognitive Decline Rates

To test if CR values correlate with cognitive decline rates, Pearson correlations were computed between the two. Brain regions with significant correlations (after correction) were marked. Additionally, the sum of CR values across all brain regions was correlated with cognitive decline rates to identify which cognitive functions are most protected by CR (Figure 1.c.②).

**Figure 1.**
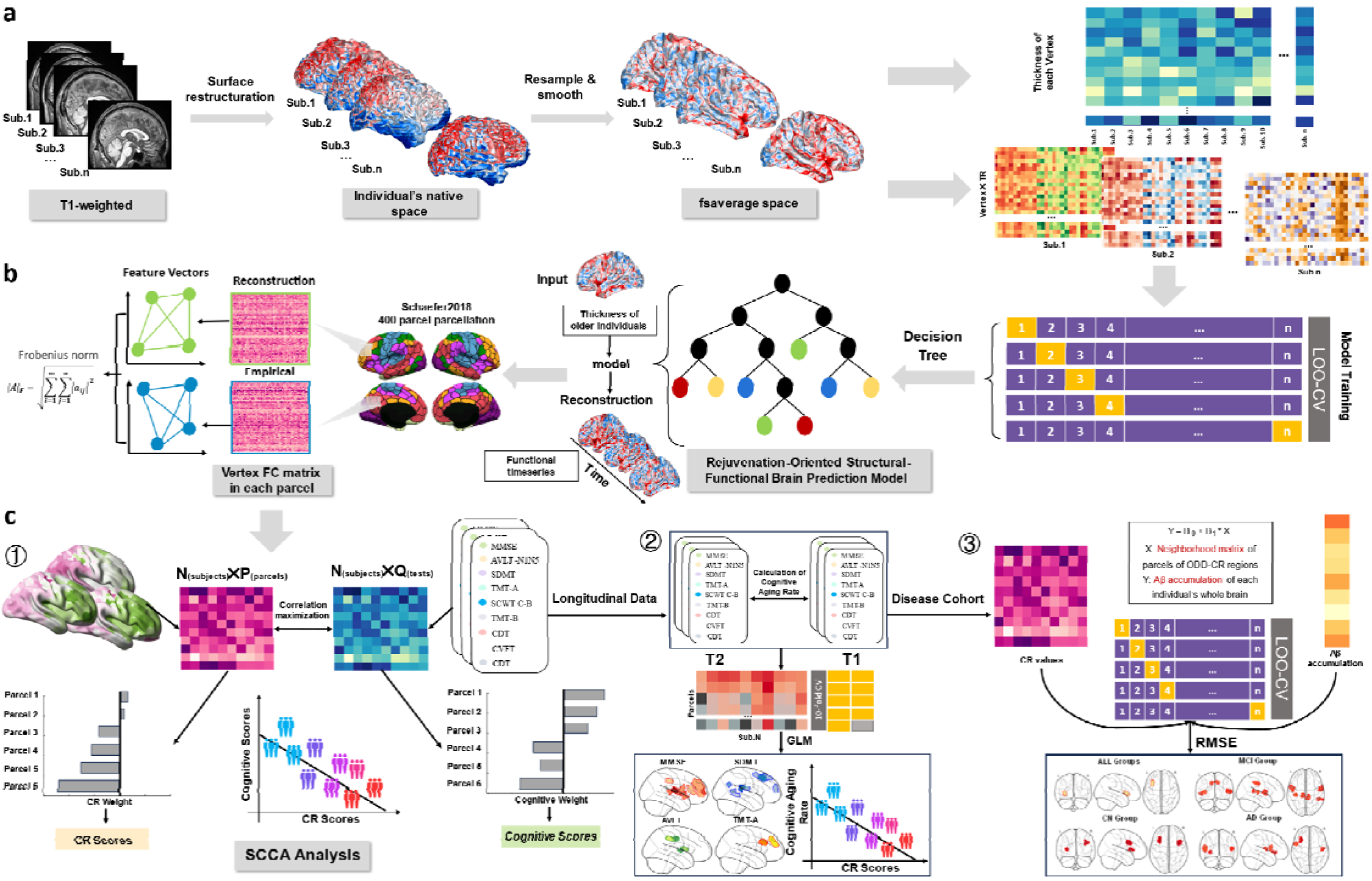
Methodological Framework for Cognitive Reserve (CR) Quantification and Analysis. (a) Multimodal Neuroimaging Data Processing Pipeline: Structural MRI data underwent cortical reconstruction and thickness mapping via FreeSurfer (fsaverage space; 163,842 vertices/hemisphere). Resting-state fMRI data were preprocessed (motion correction, bandpass filtering) and nonlinearly registered to fsaverage space, generating vertex-wise functional matrices (vertex × time × participant). (b) Machine Learning-Based CR Measurement Decision tree regression models linked vertex-level cortical thickness to functional activity patterns in young adults (LOOCV; prediction accuracy ≥60%). Model-derived functional predictions for elderly participants were compared to empirical data via Frobenius norm of connectivity matrix differences, with higher reconstruction errors indicating elevated CR. (c) Analytical Modules: ① SCCA with ℓ[ regularization was applied to identify multivariate associations between cortical reorganization (CR spatial distribution) and multi-domain cognitive performance. ② CR-Cognition Correlations: Pearson correlations between global CR values and domain-specific cognitive decline rates. ③Aβ Prediction Modeling: Leave-one-out cross-validated (LOOCV) regression identified CR hubs predicting amyloid deposition, weighted by RMSE reduction.

#### Pathological Aging Validation Dataset

##### 2.6.7 ANOVA Analysis

Firstly, we obtained the CR values for further analysis by controlling for age, sex, and education using linear regression residuals. A one-way ANOVA was performed on the residuals of CR values across brain regions for the CN, MCI, and AD groups.Regions with significant between-group differences (FDR-corrected p<0.05) were identified and further compared using Tukey’s HSD test to determine which pairs (CN-MCI, CN-AD, MCI-AD) showed significant CR differences. Additionally, correlations were assessed between CR value differences (relative to the CN group mean) in these regions and cognitive differences between the MCI/AD groups and the CN group.

##### 2.6.8 Building a CR-Based Prediction Model for Aβ Deposition

A LOOCV method was used to build a CR-based prediction model for Aβ deposition. The steps were as follows: Take the whole-brain Aβ deposition level as the target variable (Y). Include a feature matrix of CR values from each participant’s CR-related brain regions as predictor variables (X). Evaluate each CR feature’s prediction performance using RMSE. Establish a baseline model (predicting Y’s mean) and calculate its RMSE. Select CR features with RMSE below the baseline as significant predictors. Apply inverse weighting (1/RMSE) to these significant features to determine each region’s predictive power for Aβ deposition; higher values indicate greater predictive efficacy (Figure 1.c.③).

## 3 Results

### 3.1 Participant Characteristics and Cognitive Function

The study sample comprised 1,280 healthy older adults (age range: 50–89 years; mean age: 66.45 ± 7.26 years; 61.4% female) for cognitive reserve (CR) quantification. Within this cohort, 323 participants (baseline age: 50–86 years; mean: 65.74 ± 6.73 years; 61.3% female) underwent longitudinal cognitive assessments across 660 follow-up visits, with no significant demographic differences observed between this subgroup and the main cohort (independent t-tests, *p* > 0.05). The average follow-up duration was 3.19 ± 1.34 years. An independent validation cohort of 135 participants (mean age: 68.23 ± 8.09 years; 58.5% female), including cognitively normal (CN) individuals, mild cognitive impairment (MCI) patients, and Alzheimer’s disease (AD) patients, was additionally analyzed. All participants completed a standardized neuropsychological battery assessing nine cognitive domains: Mini-Mental State Examination (MMSE),

Auditory Verbal Learning Test (AVLT-N1N5), Symbol Digit Modalities Test (SDMT) and Trail Making Test Part A (TMT-A), Trail Making Test Part B (TMT-B) and Stroop Color-Word Test (SCWT C-B), Clock Drawing Test (CDT), Category Verbal Fluency Test (CVFT), Digit Span Test (DST). Detailed demographic characteristics and cognitive test performance metrics are summarized in Table 1.

**Table 1.** Characteristics and neuropsychologic test results

|  | Large-sample Dataset | Longitudinal Dataset | Pathological aging verification dataset |  |  |  |
| --- | --- | --- | --- | --- | --- | --- |
|  | Total | Total | Total | NC | MCI | AD |
|  | n = 1280 | n = 323 | n = 135 | n = 76 | n = 30 | n = 29 |
| Sex (female, %) | 786 (61.4%) | 198 (61.3%) | 79 (58.5%) | 43 (56.6%) | 15 (50%) | 21 (72.4%) |
| Age (years) | 66.45±7.26 | 65.74±6.73 | 68.23 ± 8.09 | 67.08 ± 7.63 | 68.73 ± 8.35 | 70.72 ± 8.65 |
| Education (years) | 11.49±3.14 | 11.47±2.98 | 12.60 ± 3.80 | 13.19 ± 3.56 | 12.07 ± 3.35 | 11.62 ± 4.62 |
| MMSE | 27.34±2.35 | 27.63±2.03 | 24.11 ± 6.96 | 27.58 ± 1.65 | 26.30 ± 1.56 | 12.76 ± 7.11 |
| AVLT (N1-N5) | 27.88±9.44 | 28.98±9.22 | 21.63 ± 12.20 | 29.08 ± 8.90 | 16.90 ± 8.15 | 6.46 ± 4.68 |
| SDMT | 34.31±10.77 | 34.02±10.30 | 29.25 ± 14.42 | 35.86 ± 11.50 | 25.16 ± 9.99 | 10.19 ± 8.59 |
| TMT-A | 60.23±23.03 | 59.18±21.72 | 75.77 ± 42.87 | 58.74 ± 20.43 | 86.70 ± 39.95 | 140.20± 62.69 |
| TMT-B | 172.44±70.99 | 168.46±62.61 | 179.10 ± 79.07 | 153.58 ± 50.42 | 210.61 ± 92.43 | 311.25± 89.11 |
| SCWT C-B time | 40.67±21.05 | 40.17±22.09 | 51.94±39.18 | 42.20±18.80 | 57.93±36.95 | 89.73±79.38 |
| CDT | 24.33±4.81 | 24.05±4.85 | 21.74 ± 8.62 | 25.29 ± 4.46 | 23.83 ± 5.51 | 10.07 ± 9.67 |
| CVFT | 44.61±8.99 | 45.65±8.76 | 35.77 ± 14.35 | 43.82 ± 8.65 | 34.70 ± 9.22 | 15.79 ± 10.33 |
| DST | 12.25±2.20 | 12.20±2.10 | 11.5±3.20 | 12.52±2.68 | 11.67±1.90 | 8.61±3.88 |

Note: The measured data are represented by the mean and standard deviation. AVLT: Auditory Verbal Learning Test; SCWT C-B: Stroop Color and Word Test
C minus Stroop Color and Word Test B (time); TMT: Trail Making Test; CDT: Clock-Drawing Test; CVFT: Category Verbal Fluency Test, including fruit, vegetable, and animal fluency test; SDMT: Symbol Digit Modalities Test; DST: Digit Span Test.

### 3.2 Spatially Consistent Cognitive Reserve Networks with Dominant High-Order Functional Contributions

Using CR quantification methods outlined in the Statistical Analysis section, we mapped the whole-brain spatial distribution of CR values by averaging individual-level data across the aging cohort. As shown in Figure 2.a, CR exhibited pronounced spatial heterogeneity, characterized by a “anterior reserve-posterior decline” pattern. Specifically, CR-rich regions clustered in the medial/lateral prefrontal cortex, superior temporal gyrus, pre-/postcentral gyri, cingulate cortex, and select parietal areas (e.g., precuneus). To assess the generalizability of this spatial pattern, we further constructed a population frequency map (Figure 2.b), representing the proportion of individuals (0–100%) in which each brain region was classified as a CR hub. Higher frequency values indicated greater spatial consistency of CR hubs across the population. Correlation analysis between regional CR values and hub frequency (Figure 2.c) revealed strong positive associations (cluster-level *p* < 0.05, FDR-corrected), confirming that CR-rich regions overlapped significantly with high-frequency hubs. By intersecting regions with above-average CR values (top 30%) and high hub frequency (>70%), we identified spatially consistent “core cognitive reserve regions” in healthy older adults (Figure 2.d).

**Figure 2.**
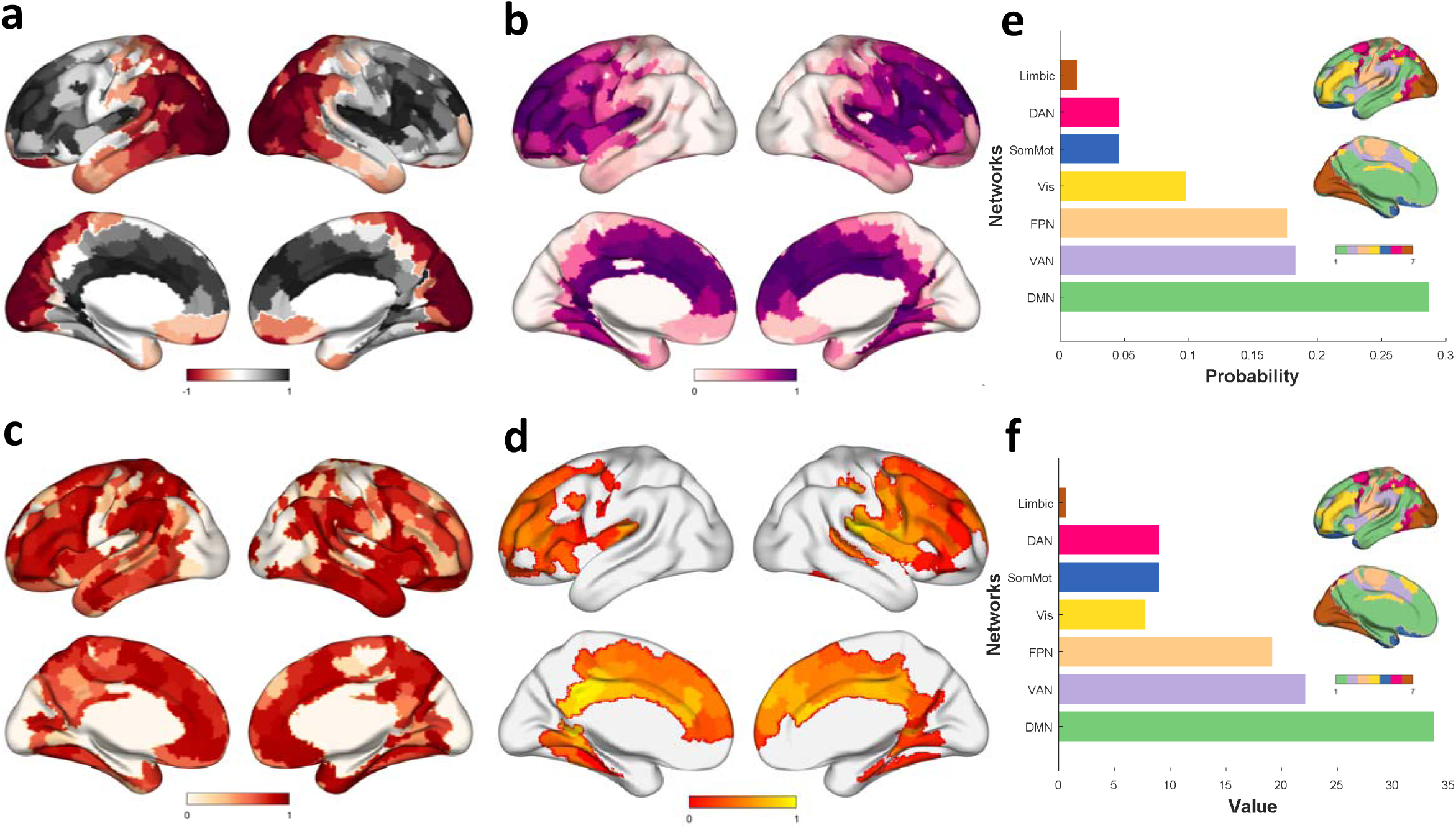
Spatial Distribution and Functional Architecture of Core Cognitive Reserve Regions. (a) Whole-brain spatial distribution of cognitive reserve (CR) values, showing an anterior-posterior gradient with enriched CR in prefrontal, temporal, cingulate, and parietal regions; (b) Population frequency map highlighting brain regions consistently classified as CR hubs across >70% of individuals; (c) Positive correlation (p < 0.05, FDR-corrected) between regional CR values and hub frequency, validating overlap between CR-rich areas and high-frequency hubs. (d) Spatially consistent “core CR regions” identified via intersection of top 30% CR values and >70% hub frequency. (e) Functional network decomposition reveals dominance of Default Mode (DMN), Ventral Attention (VAN), and Frontoparietal (FPN) networks in CR hubs (>65%). (f) Ranking by total CR magnitude reinforces DMN/VAN/FPN’s role in cognitive resilience.

To delineate the functional architecture underlying these core regions, we parsed their composition across seven canonical functional networks (Figure 2.e). After normalizing for network size (i.e., parcel count), the Default Mode Network (DMN), Ventral Attention Network (VAN), and Frontoparietal Network (FPN) dominated CR hubs, collectively accounting for >65% of all CR-rich regions. These networks are known for their high functional gradients and roles in integrative cognitive processes. Furthermore, ranking networks by total CR magnitude (summed CR values within each network) reaffirmed the prominence of DMN, VAN, and FPN (Figure 2.f), suggesting their critical involvement in sustaining cognitive resilience through adaptive functional mechanisms.

### 3.3 Age- and Sex-Stratified Differences in CR regions

We examined the impact of age, sex, and education on CR values within reserve regions using covariate-adjusted residuals. Participants were stratified into younger-old (age ≤65 years) and older-old (age >65 years) groups. Age exerted significant effects: younger-old adults exhibited higher CR values in the medial/lateral prefrontal cortex, middle frontal gyrus, and pre-/postcentral gyri, while older-old adults showed elevated CR in the fusiform gyrus, lingual gyrus, parahippocampal regions, and middle temporal gyrus (Figure 3.a). Sex differences were also pronounced: females demonstrated higher CR in the cingulate cortex, precuneus, superior frontal gyrus, and precentral gyrus, whereas males exhibited higher CR in the middle/superior frontal gyrus, superior temporal gyrus, fusiform gyrus, and parahippocampal regions (Figure 3.b). No significant CR differences were observed between high-education (≥14 years) and low-education groups.

**Figure 3.**
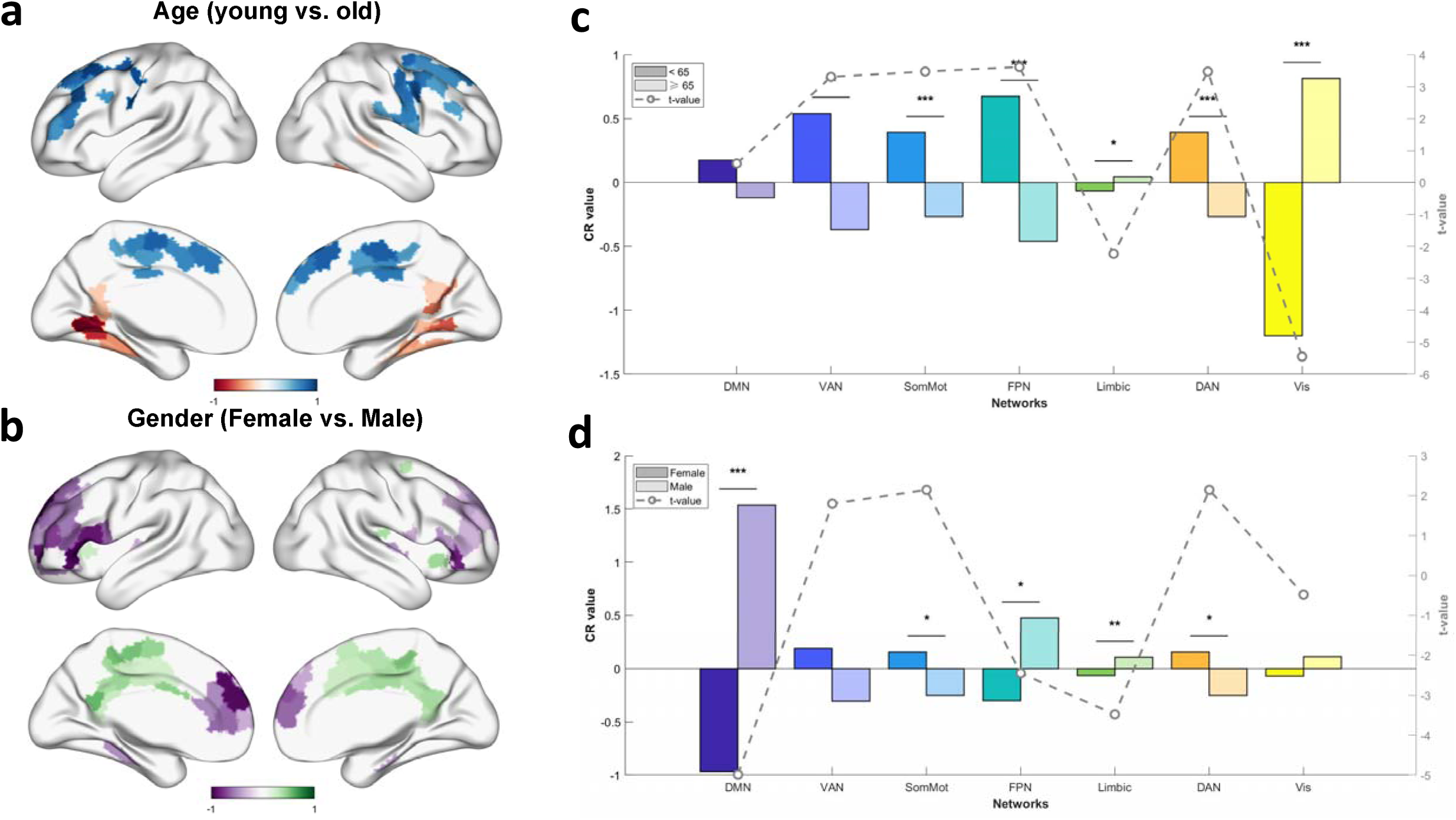
Age and Sex Differences in Cognitive Reserve Networks. (a) Younger-old adults (< 65y) show higher CR in prefrontal and sensorimotor regions (blue), while older-old adults (≥ 65y) exhibit elevated CR in temporal and parahippocampal areas (red); (b) Females display higher CR in cingulate-precuneus and frontal regions (green), males in frontotemporal and parahippocampal areas (purple); (c) Network-level age differences in CR. Younger-old adults displayed higher CR in the ventral attention (VAN), somatomotor (SomMot), frontoparietal (FPN), and dorsal attention (DAN) networks, but lower CR in visual (Vis) and limbic networks compared to older-old adults. Bar heights represent normalized CR values; (d) Network-level sex differences in CR. Males showed higher CR in the default mode (DMN), FPN, and limbic networks, while females exhibited higher CR in SomMot and DAN networks. Networks are ranked by effect size.

Further analysis of network-level CR differences revealed that younger-old adults displayed higher CR in the ventral attention (VAN), somatomotor (SomMot), frontoparietal network (FPN), and dorsal attention (DAN) networks but lower CR in visual (Vis) and limbic networks compared to older-old adults (Figure 3.c). Males exhibited higher CR in the default mode (DMN), FPN, and limbic networks, while females showed higher CR in SomMot and DAN networks (Figure 3.d). These findings underscore age- and sex-dependent reconfiguration of CR networks, with distinct functional domains preferentially contributing to cognitive resilience across demographic subgroups.

### 3.4 Domain-Specific and Cross-Phenotype Integration of CR regions Predict Cognitive performance

Using sparse canonical correlation analysis (SCCA), we identified four robust multivariate patterns linking CR spatial distributions to distinct cognitive phenotypes (processing speed, episodic memory, cognitive control, working memory). In the training dataset, these patterns exhibited significant associations (canonical r = 0.15–0.23, Bonferroni-corrected p < 0.05), which were replicated in the test dataset (r = 0.10–0.20). Figure 4.b shows the contribution weights of neuropsychological tests in the four cognitive phenotypes. Each phenotype mapped to spatially distinct CR hubs (Figure 4a). For instance, the reserve regions for processing speed phenotypes are mainly in the right supramarginal gyrus, middle cingulate gyrus, and left precuneus. For episodic memory phenotypes, they are focused in the bilateral superior frontal gyrus, left middle cingulate gyrus, and medial superior frontal gyrus. Cognitive control related regions lie in the inferior orbital frontal and anterior cingulate regions, and those for working memory phenotypes are related in the right middle temporal gyrus and precuneus.

**Figure 4.**
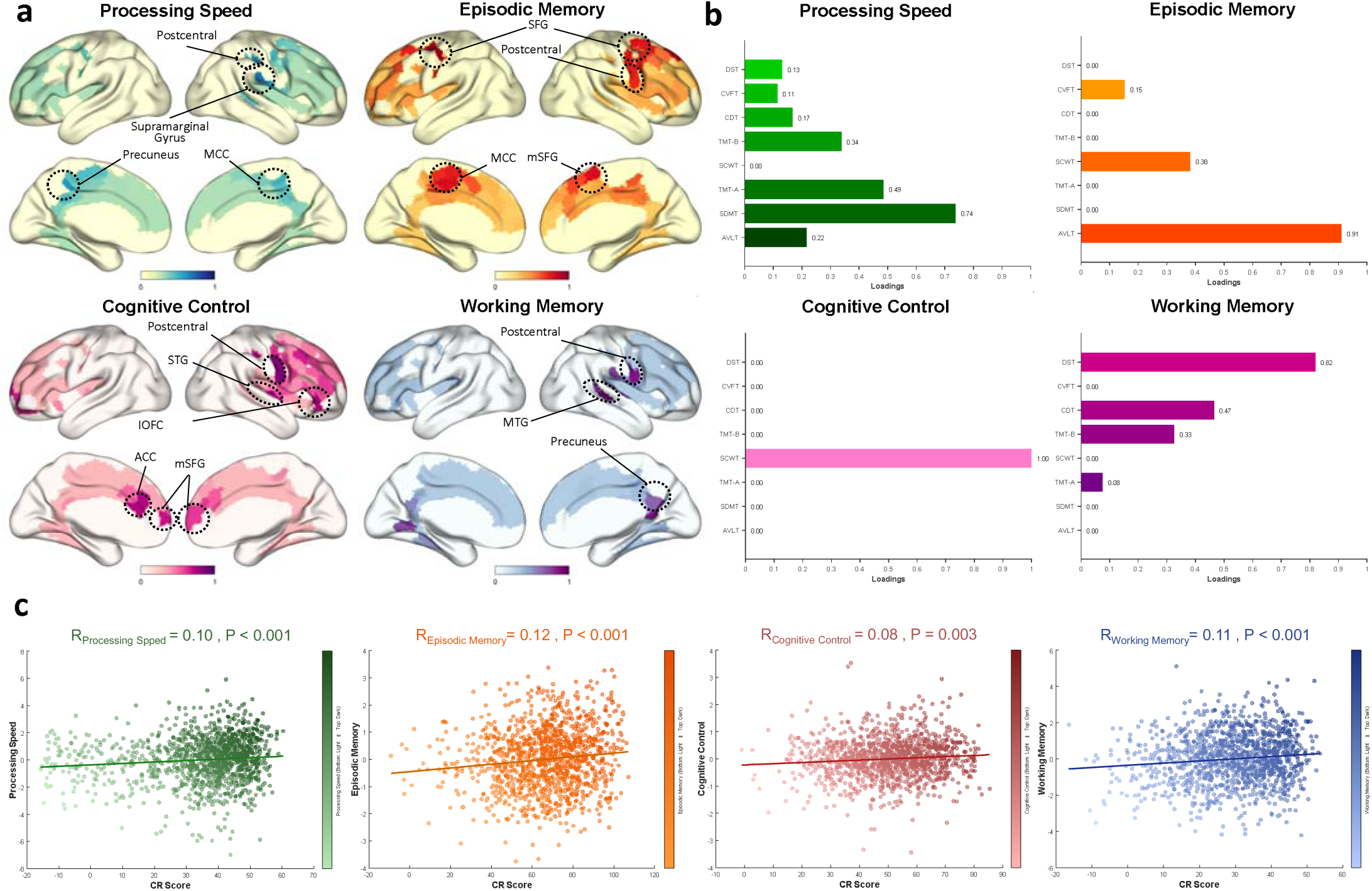
Domain-Specific and Cross-Phenotype Integration of Cognitive Reserve (CR) Networks. (a) Spatial maps of CR hubs linked to distinct cognitive phenotypes. Processing speed: right supramarginal gyrus (SMG), middle cingulate (MCC), left precuneus (PCUN); episodic memory: bilateral superior frontal gyrus (SFG), left MCC, medial SFG; cognitive control: inferior orbital frontal cortex (IOFC), anterior cingulate (ACC); working memory: right middle temporal gyrus (MTG), PCUN. The postcentral gyrus (PoCG) and SFG contributed to all phenotypes (shared hubs, yellow outlines); (b) Contribution weights of neuropsychological tests to four cognitive phenotypes (processing speed, episodic memory, cognitive control, working memory) derived from sparse canonical correlation analysis (SCCA); (c) Correlation between summed CR values of phenotype-specific hubs and composite cognitive scores (weighted by neuropsychological test contributions). Episodic memory and working memory showed the strongest associations.

Notably, the postcentral gyrus and superior frontal gyrus contributed to all four phenotypes, while the middle cingulate and precuneus supported multiple phenotypes (Figure 4b), suggesting that CR hubs form flexible subsystems through combinatorial arrangements to support diverse cognitive demands.

Furthermore, the summed CR values of phenotype-specific hubs strongly correlated with composite cognitive scores (weighted by neuropsychological test contributions). This relationship was most pronounced for episodic memory and working memory (Figure 4c), indicating that higher CR in targeted networks predicts superior domain-specific performance.

### 3.5 CR Networks Predict Multi-Domain Attenuation of Cognitive Decline in Aging

To validate whether elevated CR values in reserve regions correlate not only with better baseline cognitive performance but also with slower longitudinal cognitive decline, we calculated annualized cognitive decline rates (see Methods) and examined their associations with regional CR values. Spatial mapping revealed distinct CR hubs significantly linked to reduced decline rates across cognitive domains (Figure 5.a & c). The most extensive CR-protective effects were observed for general cognition (MMSE) and working memory (DST), followed by visuospatial (CDT) and language (VFT) domains. In contrast, episodic memory (AVLT), processing speed (TMT-A), and cognitive control (SCWT) exhibited narrower, phenotype-specific CR associations.

**Figure 5.**
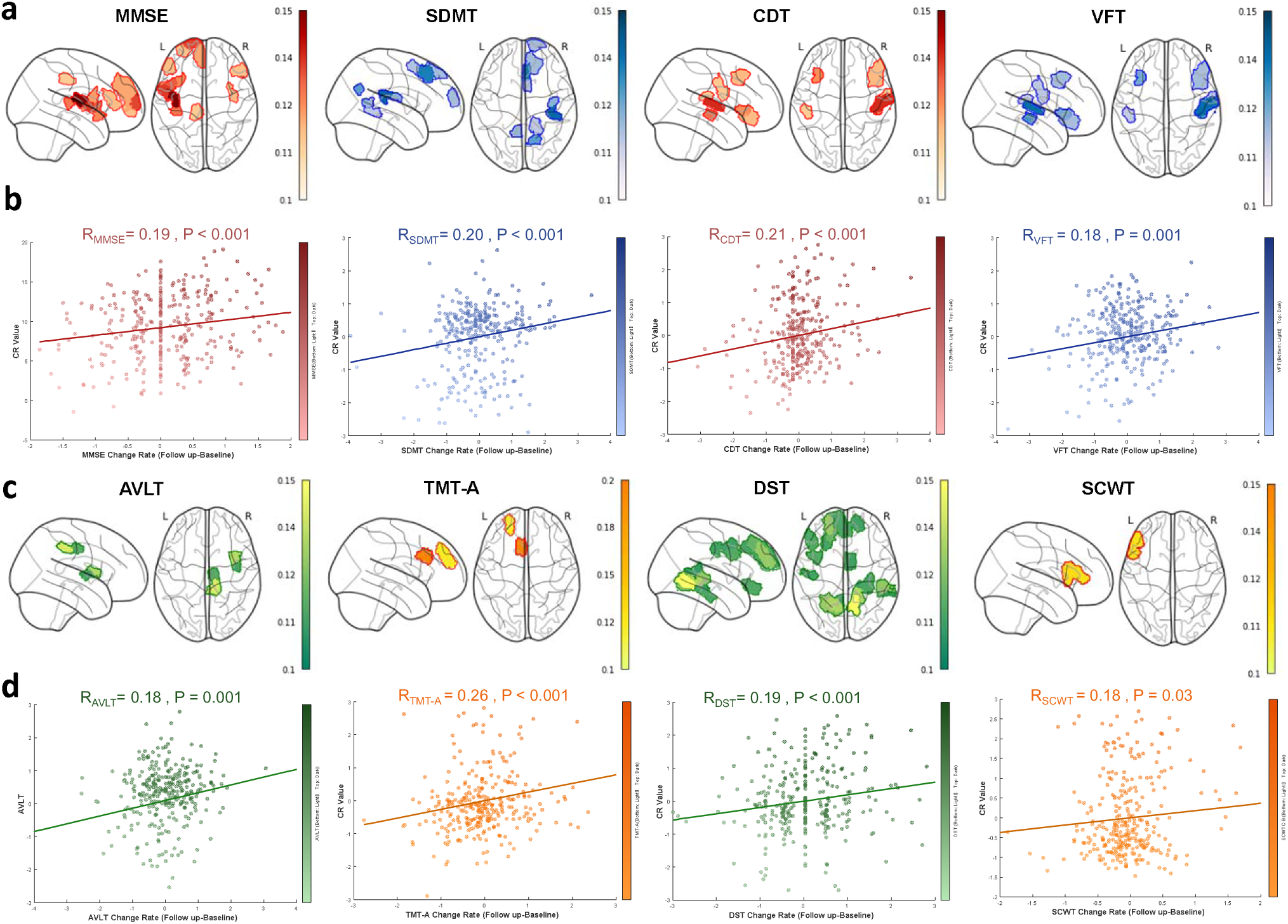
Cognitive Reserve (CR) Networks Predict Slowed Cognitive Decline in Aging. (a & c) Spatial distribution of CR hubs associated with reduced decline rates across cognitive domains. The strongest protective effects (warmer colors) were observed for general cognition (MMSE) and working memory (DST), followed by visuospatial (CDT) and language (VFT) domains. Episodic memory (AVLT), processing speed (TMT-A), and cognitive control (SCWT) showed narrower, phenotype-specific associations; (b & c) Negative correlation between aggregated CR values (summed across decline-related hubs) and global cognitive decline rates. Higher CR predicts slower longitudinal cognitive deterioration, supporting CR as a neuroprotective biomarker against aging-related decline.

Aggregating CR values across decline-related hubs demonstrated a robust correlation between total CR and global cognitive alternation rates (Figure 5.b & d), indicating that higher CR predicts slower aging-related cognitive deterioration. These findings position CR as a quantifiable neuroprotective biomarker, aligning with theoretical frameworks of cognitive reserve that emphasize adaptive resilience against age- and pathology-driven functional decline.

### 3.6 CR value Reveal Disease Stage-Specific Vulnerability

Comparative analysis of CR distributions across cognitively normal (CN), mild cognitive impairment (MCI), and Alzheimer’s disease (AD) groups revealed distinct neurobiological signatures of pathological aging. While spatial CR patterns in CN and pathological groups (MCI/AD) showed no global differences (Figure 6.a), summed CR values across reserve regions were significantly higher in CN compared to MCI and AD (Figure 6.b), with no MCI-AD group differences. Network-level analyses of covariate-adjusted CR residuals demonstrated that MCI exhibited pronounced CR reductions in the default mode (DMN), ventral attention (VAN), frontoparietal (FPN), and limbic networks relative to CN, whereas AD showed CR deficits only in VAN (Figures 6.c).

**Figure 6.**
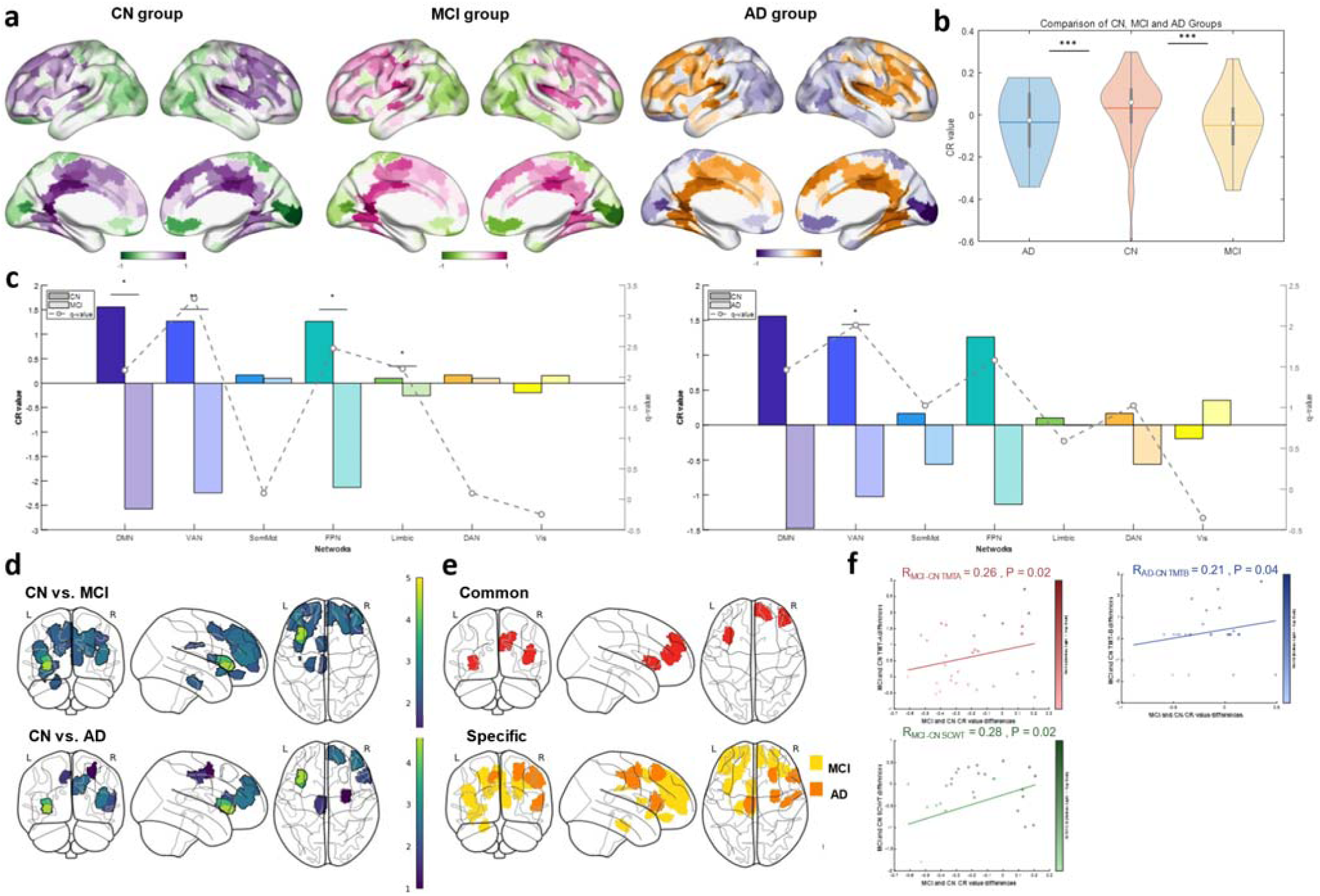
Stage-Specific Vulnerability of Cognitive Reserve (CR) in Pathological Aging. (a) Whole-brain spatial CR patterns in cognitively normal (CN), mild cognitive impairment (MCI), and Alzheimer’s disease (AD) groups. No global differences were observed between CN and pathological groups (MCI/AD); (b) Summed CR values across reserve regions. CN exhibited significantly higher CR than MCI and AD (**p<0.001), with no MCI-AD differences; (c) Network-level CR differences. MCI showed reduced CR in default mode (DMN), ventral attention (VAN), frontoparietal (FPN), and limbic networks vs. CN, while AD deficits were restricted to VAN. Bars: mean ± SEM (\**p*<0.05, \*\**p*<0.01, ANOVA with Tukey HSD). (d) Spatial maps of CR reductions in MCI (vs. CN) and AD (vs. CN). MCI exhibited broader deficits in prefrontal and temporal hubs (lighter colors); (e) Overlap and specificity of CR-deficient regions. MCI-specific reductions dominated in anterior prefrontal and temporal regions (orange), while shared MCI-AD deficits were minimal (purple); (f) CR deficits in MCI/AD correlated with domain-specific cognitive decline (e.g., episodic memory [AVLT], executive function [SCWT]; *p*<0.05).

Spatial mapping of group differences (Figures 7.d) highlighted broader CR reductions in MCI compared to AD, particularly in anterior medial/lateral prefrontal and temporal regions—key hubs of cognitive reserve (Figures 6.e). Notably, CR differences in overlapping regions (e.g., superior frontal gyrus) were minimal between MCI and AD, suggesting MCI-specific vulnerability preceding widespread AD pathology. Furthermore, CR deficits in MCI/AD significantly correlated with domain-specific cognitive decline (e.g., memory, executive function; Figure 6.f), reinforcing CR’s role as a mediator of clinical symptom severity.

**Figure 7.**
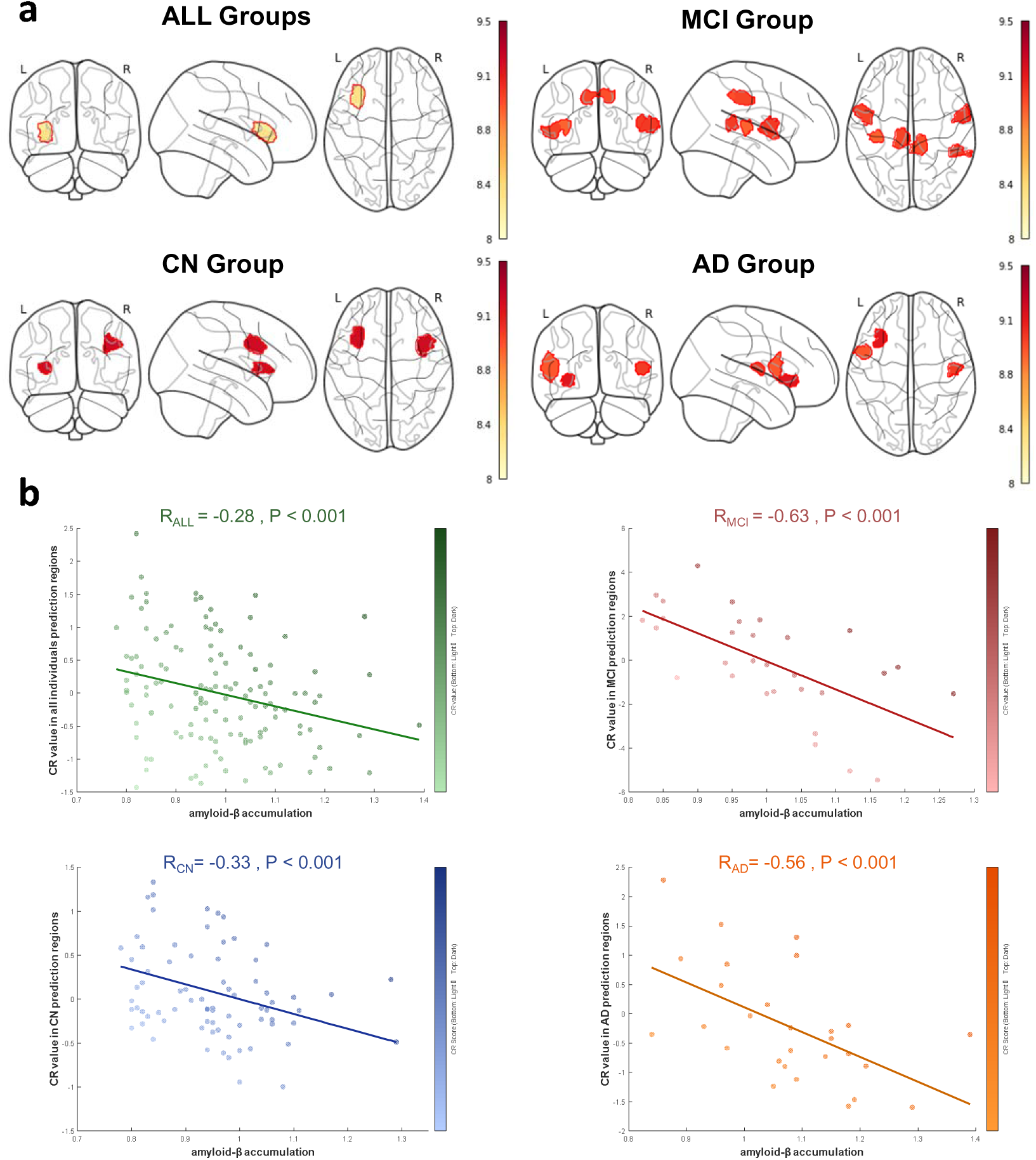
Stage-Specific Predictive Efficacy of CR Networks for Aβ Deposition. (a) Regional predictive power of CR hubs for Aβ deposition across CN, MCI, and AD groups. In the combined cohort and CN group, only the inferior orbital frontal cortex (IOFC) showed modest associations. AD exhibited predictive CR hubs in the Rolandic operculum (RO) and precentral gyrus (PreCG), while MCI demonstrated the highest sensitivity with hubs in the superior/middle temporal gyri (STG/MTG), middle cingulate (MCC), precuneus (PCUN), and Heschl’s gyrus; (b) Inverse correlation between aggregated CR values of top-predictive hubs and global Aβ burden (SUVR). The strongest association was observed in MCI, indicating CR’s neuroprotective role against amyloid accumulation during preclinical stages.

These findings identify MCI as a critical window of CR network sensitivity, offering early biomarkers for pathological progression, while underscoring the divergent neuroadaptive trajectories between MCI and AD.

### 3.7 CR Networks Predict Stage-Specific Aβ Deposition

Using a leave-one-out cross-validated generalized linear regression framework (see Methods), we evaluated the ability of CR values in reserve regions to predict whole-brain Aβ deposition across cognitively normal (CN), mild cognitive impairment (MCI), and Alzheimer’s disease (AD) groups. In the combined cohort and CN group, CR hubs exhibited limited predictive sensitivity, with only the inferior orbital frontal cortex showing modest associations (Figure 7.a). In AD, predictive efficacy extended to the Rolandic operculum and precentral gyrus, while MCI demonstrated the highest sensitivity, with CR hubs in the superior/middle temporal gyri, middle cingulate, precuneus, and Heschl’s gyrus robustly predicting Aβ deposition.

Critically, aggregated CR values from top-predictive hubs inversely correlated with global Aβ burden (Figure 7.b), indicating that higher CR is associated with reduced amyloid pathology. This relationship was strongest in MCI, suggesting CR networks act as early neuroprotective buffers against amyloid accumulation during the preclinical phase.

## 4 Discussion

This study established a novel metric for quantifying cognitive reserve (CR), integrating brain functional signatures that best capture CR characteristics with structural underpinnings to elucidate its neurobiological mechanisms. The spatial distribution of CR values across a large aging cohort revealed core reserve hubs concentrated in the prefrontal cortex, cingulate gyrus, and precuneus, predominantly organized within high-order functional networks—including the default mode network (DMN), ventral attention network (VAN), and frontoparietal network (FPN). Domain-specific cognitive phenotypes (processing speed, episodic memory, cognitive control, working memory) exhibited distinct spatial associations with CR networks, where key regions (e.g., postcentral gyrus, superior frontal gyrus, middle cingulate) formed adaptive subsystems to support cognition through combinatorial interactions. Longitudinal analyses demonstrated significant negative correlations between CR values and rates of cognitive decline, indicating that elevated CR predicts attenuated aging-related cognitive deterioration. Pathological aging cohorts (MCI and AD) showed markedly reduced CR compared to cognitively normal (CN) individuals, with MCI exhibiting greater sensitivity to CR alterations than AD, particularly in prefrontal and temporal hubs. Critically, CR deficits were robustly associated with increased Aβ deposition, highlighting its potential as an early biomarker for preclinical amyloid pathology. These findings position CR as a multifaceted neuroprotective framework, bridging functional resilience, structural integrity, and pathological progression, while offering actionable targets for interventions aimed at mitigating cognitive decline in aging and neurodegeneration.

This study introduces a novel cognitive reserve (CR) metric combining functional and structural brain features to map CR mechanisms. Core CR hubs localized to prefrontal, cingulate, and precuneus regions, primarily within high-order networks (DMN, VAN, FPN). Domain-specific cognitive phenotypes (e.g., memory, attention) linked to distinct CR networks, with adaptive subsystems (e.g., postcentral/superior frontal gyri) supporting cognition. Higher CR predicted slower cognitive decline. Pathological aging (MCI/AD) showed reduced CR, with MCI exhibiting heightened sensitivity in prefrontal/temporal regions. CR inversely correlated with Aβ deposition, marking it as an early biomarker. CR emerges as a multidimensional neuroprotective system, integrating resilience, structure, and pathology, guiding interventions against cognitive decline.

The pronounced heterogeneity in cognitive aging presents a critical challenge for unraveling its mechanisms and mitigating aging-related decline. Cognitive reserve (CR) serves as a pivotal concept in elucidating this phenomenon [55–57]. From a lifespan development perspective, individual differences in cognitive aging manifest primarily in peak cognitive performance during early adulthood and the subsequent rate of decline with advancing age [58, 59]. Traditional CR proxies, such as education and occupation, emphasize pre-aging advantages in neurobiological resources [60]. However, contemporary research increasingly focuses on adaptive strategies employed during aging, positing that CR modulates the interplay between brain state and clinical pathology, thereby altering cognitive-behavioral correlations under equivalent structural integrity [5]. Prior studies demonstrate that CR modifies the relationship between brain structure and cognition: individuals with higher CR exhibit superior cognitive performance despite comparable gray matter volume or white matter integrity [61–63], suggesting enhanced utilization of available neural resources. Furthermore, elevated CR is associated with preserved network efficiency even amid gray matter atrophy [19]. Our novel CR metric advances prior frameworks by directly quantifying neurobiological reserve, capturing older adults’ capacity to flexibly adapt to developmental changes. Strikingly, we identified “reserve hubs”—brain regions where functional activity in older adults exceeds the maximal threshold predicted by youthful structural models. These hubs exhibit high inter-individual spatial consistency, yet the magnitude of functional excess varies systematically, reflecting individual differences in CR. This divergence underscores CR’s role in buffering age-related neural decline while highlighting its potential as a biomarker for personalized cognitive resilience strategies.

Notably, the identified reserve hubs—localized to the medial/lateral frontal cortex[64–66], cingulate cortex[67, 68], and precuneus[69, 70]—reflect intrinsic neurobiological mechanisms underlying cognitive reserve. The CR regions, predominantly organized within high-order functional networks (default mode network [DMN], ventral attention network [VAN], and frontoparietal network [FPN]), occupy apex positions along functional gradients and serve as integrative hubs for cross-modal information processing [71]. Longitudinal studies of cortical atrophy reveal divergent trajectories between normal and pathological aging: typical aging exhibits relatively uniform atrophy across frontal and temporal regions, whereas pathological aging (e.g., Alzheimer’s disease [AD]) is characterized by accelerated atrophy focused in the temporal lobe, with relative sparing of prefrontal regions [72]. Further analyses of low-risk populations (Aβ-negative, APOE ε4 non-carriers) demonstrate that mild cognitive impairment (MCI) and AD cohorts exhibit maximal standardized atrophy rates in temporal regions, while low-risk individuals retain a frontal-temporal balanced atrophy pattern resembling normal aging [73, 74].

According to the multiple factor framework of brain aging [75, 76], pathological cognitive aging (PCA) and normal cognitive aging (NCA) follow distinct trajectories: PCA is driven by early medial temporal lobe vulnerability and rapid pathological spread, whereas NCA predominantly involves gradual decline in fronto-striatal circuitry. Critically, our reserve hubs overlap extensively with fronto-striatal networks, suggesting that CR variations in these hubs are critical markers of individual differences in developmental trajectories among the NCA cohort [77]. Individuals failing to achieve NCA-level CR in these hubs may face elevated risks of transitioning to PCA, highlighting CR’s dual role as both a biomarker of resilience and a predictor of pathological progression.

Cognitive reserve (CR), as a neurobiological property of the brain, enables individuals to achieve better-than-expected cognitive performance despite age-related neural decline by leveraging existing neurocognitive resources. Neuroplasticity—the brain’s ability to optimize neural circuit efficiency and adapt to internal and external demands—provides a physiological foundation for CR [3]. In the context of memory aging, neuroplastic mechanisms operate through three primary pathways: **neurogenesis** (generation of new neurons), **structural remodeling** (strengthened connectivity, volumetric expansion, or reorganization of existing pathways), and **functional engagement** (adaptive reconfiguration of activation patterns) [78]. While neurogenesis and structural remodeling are resource-intensive and decline with aging, functional engagement emerges as a dominant adaptive strategy. The **functional plasticity hypothesis of cognitive aging** posits that age-related structural atrophy disrupts cognition by impairing region-dependent processes, yet the aging brain compensates by recruiting alternative regions (e.g., prefrontal and parietal cortices) through strategic allocation and functional reorganization [79–81]. Our findings extend this framework by revealing a novel form of functional plasticity: older adults exhibit functional performance exceeding structural capacity thresholds in regions such as the prefrontal cortex, even in areas with notable atrophy. Remarkably, these regions maintain functional integrity despite structural degeneration, diverging from classical compensatory mechanisms, which typically involve hyperactivation of intact regions to enhance task performance [82]. Critically, our CR metric captures this dissociation between structural decline and preserved function, highlighting a mechanism distinct from traditional compensation. This phenomenon parallels a “software upgrade” compensating for aging “hardware,” where functional optimization offsets structural limitations. By quantifying CR as the gap between predicted and observed functional performance, our framework identifies neuroplastic buffers that sustain cognition despite atrophy. These insights inform targeted interventions to amplify CR-driven plasticity, offering biomarkers to evaluate therapeutic efficacy and guide personalized strategies to mitigate cognitive decline.

Educational attainment, a widely studied proxy for cognitive reserve (CR), is recognized as a robust protective factor against age-related cognitive decline [83]. However, in this study, no significant differences in CR values were observed between high-education (≥14 years) and low-education groups within reserve hubs. We posit that this discrepancy arises from divergent mechanistic emphases: education primarily reflects early-life advantages in neurobiological “hardware” (e.g., structural reserve), whereas our CR metric captures adaptive “software” mechanisms that optimize functional performance within existing structural constraints. Extensive evidence demonstrates that higher educational attainment confers broad protective effects on late-life cognition, including executive function, visuospatial processing, language, and working memory [84, 85]. It also reduces Alzheimer’s disease and dementia risk [86–88], with stronger benefits for attention and executive function in preclinical stages compared to dementia cohorts [89]. Critically, however, education’s protective effects are attributed to elevated peak cognitive capacity in early adulthood rather than slowed decline rates [90, 91]. Thus, while education helps maintain baseline cognitive performance in normal aging (NCA), it may inadequately buffer against age- or pathology-driven cognitive deterioration. This distinction aligns with our findings: CR, as a dynamic functional adaptation metric, requires individuals to optimize neural resource allocation (“software”) despite structural limitations (“hardware”). High education alone, without concurrent functional adaptation, fails to enhance CR. This underscores CR’s unique role in quantifying resilience through real-time neural efficiency rather than static structural reserves, offering novel insights into interventions targeting adaptive plasticity to mitigate aging-related decline.

CR hubs exhibit domain-specific and domain-general contributions to multi-domain cognition. Subregions within reserve hubs—such as the medial/lateral superior frontal gyrus, precuneus, and cingulate cortex—demonstrated both specialized and overlapping associations with distinct cognitive domains in multivariate analyses. These regions overlap extensively with the fronto-striatal circuitry, a critical network linking the prefrontal cortex and striatum to support higher-order cognitive functions through complex cross-regional interactions [92, 93]. Our findings suggest that the relationship between CR and cognition depends on the differential contributions of key subregions, with varying functional weights across domains. However, the specific mechanisms by which these subregional circuits mediate CR’s effects on cognition remain to be elucidated. Notably, while CR values in domain-specific hubs showed statistically significant correlations with cognitive scores, the effect sizes were modest (r < 0.2), raising questions about CR’s robustness as a standalone metric for quantifying cognitive heterogeneity. This limitation, however, aligns with our operational definition of CR as a dynamic buffer against decline rather than a direct correlate of baseline cognitive performance [2, 4, 6]. Longitudinal analyses strengthened CR’s validity: higher CR values predicted slower cognitive decline, independent of education—a proxy emphasizing early-life structural advantages. This dissociation underscores CR’s unique utility in capturing adaptive resilience mechanisms that mitigate aging-related deterioration, positioning it as a complementary biomarker to traditional structural reserve metrics.

The concept of cognitive reserve (CR) extends beyond mitigating age-related cognitive decline to critically regulating the risk of transitioning from normal cognitive aging (NCA) to pathological cognitive aging (PCA). Unlike conventional biomarkers of aging progression (e.g., gray matter volume, cortical thickness), CR values do not exhibit a continuum across cognitively normal (CN), mild cognitive impairment (MCI), and Alzheimer’s disease (AD) groups. Instead, MCI shows broader CR reductions in reserve hubs and heightened sensitivity to CR alterations compared to AD. This divergence stems from stage-specific neuroplastic capacity: MCI, as an early pathological stage, retains partial neuroplasticity to adapt to functional deficits through compensatory mechanisms, whereas AD exhibits pervasive failure of such adaptation.

According to the Exploration-Selection-Refinement Model [94], cognitive processing relies on a dynamic equilibrium between task demands and functional supply. In NCA, CR enables individuals to restore this equilibrium by enhancing functional supply (e.g., recruiting alternative networks) when structural decline (e.g., atrophy) reduces supply. MCI, while retaining this capacity, demonstrates inefficient CR utilization, marked by pronounced CR reductions in the medial/lateral prefrontal cortex—key hubs for adaptive resource allocation. In contrast, AD’s widespread structural damage disrupts the “hardware” necessary for CR-driven “software” optimization, rendering CR metrics unreliable in these regions. Notably, in MCI, higher CR in the superior temporal gyrus and precuneus strongly predicts lower Aβ deposition, positioning CR as a biomarker for preclinical AD detection. This suggests that early interventions boosting CR, potentially through cognitive training or neuromodulation, could enhance functional plasticity, delay amyloid accumulation, and decelerate pathological progression. These findings redefine CR as a dynamic gatekeeper of neuroplastic resilience, offering a mechanistic framework to stratify NCA-to-PCA transition risk and guide precision interventions.

## 5 Limitations and Future Directions

The current study has two primary limitations that warrant consideration. First, the absence of longitudinal neuroimaging data in pathological aging cohorts restricts our ability to delineate how CR dynamically mitigates pathological progression or modulates the transition risk from normal to pathological aging. While longitudinal cognitive assessments were available for a subset of participants (n=323), the limited temporal scope (mean follow-up: 3.19 ±1.34 years) and partial behavioral coverage constrain causal inferences between CR and cognitive decline. Extended follow-up studies incorporating serial neuroimaging (e.g., annual MRI/PET) are critical to validate CR’s temporal neuroprotective effects and unravel bidirectional relationships between CR, structural atrophy, and pathological spread.

Second, the exclusion of complementary biomarkers, such as tau pathology (e.g., tau-PET) or cerebral metabolism (e.g., FDG-PET), limits the biological interpretability of CR mechanisms. Future studies should integrate multi-modal neuroimaging to disentangle how CR interacts with hallmark Alzheimer’s pathologies (Aβ/tau) and metabolic dysregulation.

Addressing these gaps will advance CR from a theoretical construct to a multidimensional biomarker system, bridging molecular, structural, functional, and behavioral layers of cognitive resilience. This integrative approach is essential for developing precision interventions that target CR-specific pathways to delay or prevent pathological aging.

## 6 Conclusion

This study delineates cognitive reserve (CR) as a spatially organized, high-order functional network phenomenon anchored in the default mode, ventral attention, and frontoparietal systems. CR hubs exhibit age- and sex-dependent reconfigurations, predict domain-specific cognitive resilience against decline, and demonstrate stage-specific associations with Aβ deposition, most prominently in mild cognitive impairment. These findings position CR mapping as a clinically actionable biomarker for early neurodegenerative detection and personalized cognitive preservation strategies, bridging neuroadaptive plasticity with pathological progression in aging.

## Acknowledgments and Funding Disclosure

This work was supported by STI2030-Major Projects (2022ZD0211600), the Natural Science Foundation of China (Grants No. 32171085), the National Natural Science Foundation of China(Grants No. 12405062), State Key Program of National Natural Science of China (Grants No. 82130118), the Fundamental Research Funds for the Central Public welfare research institution (ZZ13-YQ-073, Z0601), Open Research Fund of the State Key Laboratory of Cognitive Neuroscience and Learning and Tang Scholar.

## Authors’ Contributions

LYM and LX conceived the study. LYM and ZXY participated in the data collection. LYM carried out statistical analysis. LYM analyzed and interpreted the data. LYM wrote the manuscript. LX revised the manuscript. ZZJ supervised and coordinated the study. All authors contributed to the improvement of this manuscript and approved the final version for submission.

## Data availability

Raw data of the older adults with their completed narrative and extensive neuropsychological tests and MRI scans are available from the corresponding author on reasonable request. Restriction of raw data is to protect the privacy of participants. Source data are provided with this paper. Source data are provided with this paper.

## Code availability

Custom codes are variable at https://github.com/Rainmon2020/Cognitive-Reserve/upload.

## Reference

[1] Lindenberger U. Human cognitive aging: corriger la fortune? Science. 2014;346:572–8.

[2] Stern Y, Arenaza-Urquijo EM, Bartres-Faz D, Belleville S, Cantilon M, Chetelat G, et al. Whitepaper: Defining and investigating cognitive reserve, brain reserve, and brain maintenance. Alzheimers Dement. 2020;16:1305–11.

[3] Barulli D, Stern Y. Efficiency, capacity, compensation, maintenance, plasticity: emerging concepts in cognitive reserve. Trends Cogn Sci. 2013;17:502–9.

[4] Stern Y, Albert M, Barnes CA, Cabeza R, Pascual-Leone A, Rapp PR. A framework for concepts of reserve and resilience in aging. Neurobiol Aging. 2023;124:100–3.

[5] Stern Y. An approach to studying the neural correlates of reserve. Brain Imaging Behav. 2017;11:410–6.

[6] Stern Y, Chetelat G, Habeck C, Arenaza-Urquijo EM, Vemuri P, Estanga A, et al. Mechanisms underlying resilience in ageing. Nat Rev Neurosci. 2019;20:246.

[7] Hertzog C, Dunlosky J, Sinclair SM. Episodic feeling-of-knowing resolution derives from the quality of original encoding. Mem Cognit. 2010;38:771–84.

[8] Mukadam N, Sommerlad A, Huntley J, Livingston G. Population attributable fractions for risk factors for dementia in low-income and middle-income countries: an analysis using cross-sectional survey data. Lancet Glob Health. 2019;7:e596–e603.

[9] Franz CE, Hatton SN, Elman JA, Warren T, Gillespie NA, Whitsel NA, et al. Lifestyle and the aging brain: interactive effects of modifiable lifestyle behaviors and cognitive ability in men from midlife to old age. Neurobiol Aging. 2021;108:80–9.

[10] Smart EL, Gow AJ, Deary IJ. Occupational complexity and lifetime cognitive abilities. Neurology. 2014;83:2285–91.

[11] Dorman GS, Boccazzi JF, Flores I, O’Neill S. Relationship between occupation and cognitive performance in patients with cognitive impairment. Preliminary study. Alzheimer’s & Dementia. 2023;19:e079330.

[12] Reed BR, Mungas D, Farias ST, Harvey D, Beckett L, Widaman K, et al. Measuring cognitive reserve based on the decomposition of episodic memory variance. Brain. 2010;133:2196–209.

[13] Bettcher BM, Gross AL, Gavett BE, Widaman KF, Fletcher E, Dowling NM, et al. Dynamic change of cognitive reserve: associations with changes in brain, cognition, and diagnosis. Neurobiol Aging. 2019;83:95–104.

[14] Zahodne LB, Manly JJ, Brickman AM, Narkhede A, Griffith EY, Guzman VA, et al. Is residual memory variance a valid method for quantifying cognitive reserve? A longitudinal application. Neuropsychologia. 2015;77:260–6.

[15] Jones RN, Manly J, Glymour MM, Rentz DM, Jefferson AL, Stern Y. Conceptual and measurement challenges in research on cognitive reserve. J Int Neuropsychol Soc. 2011;17:593–601.

[16] Bartres-Faz D, Arenaza-Urquijo EM. Structural and functional imaging correlates of cognitive and brain reserve hypotheses in healthy and pathological aging. Brain Topogr. 2011;24:340–57.

[17] Conti L, Riccitelli GC, Preziosa P, Vizzino C, Marchesi O, Rocca MA, et al. Effect of cognitive reserve on structural and functional MRI measures in healthy subjects: a multiparametric assessment. J Neurol. 2021;268:1780–91.

[18] Stern Y, Habeck C, Moeller J, Scarmeas N, Anderson KE, Hilton HJ, et al. Brain networks associated with cognitive reserve in healthy young and old adults. Cereb Cortex. 2005;15:394–402.

[19] Steffener J, Brickman AM, Rakitin BC, Gazes Y, Stern Y. The impact of age-related changes on working memory functional activity. Brain imaging and behavior. 2009;3:142–53.

[20] Medaglia JD, Pasqualetti F, Hamilton RH, Thompson-Schill SL, Bassett DS. Brain and cognitive reserve: Translation via network control theory. Neurosci Biobehav Rev. 2017;75:53–64.

[21] Wilson RS, Nag S, Boyle PA, Hizel LP, Yu L, Buchman AS, et al. Neural reserve, neuronal density in the locus ceruleus, and cognitive decline. Neurology. 2013;80:1202–8.

[22] Verkhratsky A, Zorec R. Neuroglia in cognitive reserve. Mol Psychiatry. 2024;29:3962–7.

[23] Esiri MM, Chance SA. Cognitive reserve, cortical plasticity and resistance to Alzheimer’s disease. Alzheimer’s research & therapy. 2012;4:1–8.

[24] Winkler AM, Kochunov P, Blangero J, Almasy L, Zilles K, Fox PT, et al. Cortical thickness or grey matter volume? The importance of selecting the phenotype for imaging genetics studies. Neuroimage. 2010;53:1135–46.

[25] Jirsa VK, Haken H. Field theory of electromagnetic brain activity. Physical review letters. 1996;77:960.

[26] Robinson PA, Rennie CJ, Wright JJ. Propagation and stability of waves of electrical activity in the cerebral cortex. Physical Review E. 1997;56:826.

[27] Teo PC, Sapiro G, Wandell BA. Creating connected representations of cortical gray matter for functional MRI visualization. IEEE transactions on medical imaging. 1997;16:852–63.

[28] Pang JC, Aquino KM, Oldehinkel M, Robinson PA, Fulcher BD, Breakspear M, et al. Geometric constraints on human brain function. Nature. 2023;618:566–74.

[29] Zamani Esfahlani F, Faskowitz J, Slack J, Misic B, Betzel RF. Local structure-function relationships in human brain networks across the lifespan. Nat Commun. 2022;13:2053.

[30] Park HJ, Friston K. Structural and functional brain networks: from connections to cognition. Science. 2013;342:1238411.

[31] Valenzuela MJ, Sachdev P. Brain reserve and dementia: a systematic review. Psychol Med. 2006;36:441–54.

[32] Fratiglioni L, Wang H-X. Brain reserve hypothesis in dementia. Journal of Alzheimer’s disease. 2007;12:11–22.

[33] Yang C, Li X, Zhang J, Chen Y, Li H, Wei D, et al. Early prevention of cognitive impairment in the community population: The Beijing Aging Brain Rejuvenation Initiative. Alzheimers Dement. 2021;17:1610–8.

[34] Zhang M, Katzman R, Salmon D, Jin H, Cai G, Wang Z, et al. The prevalence of dementia and Alzheimer’s disease in Shanghai, China: impact of age, gender, and education. Annals of Neurology: Official Journal of the American Neurological Association and the Child Neurology Society. 1990;27:428–37.

[35] McKhann G, Drachman D, Folstein M, Katzman R, Price D, Stadlan EM. Clinical diagnosis of Alzheimer’s disease: Report of the NINCDSLADRDA Work Group* under the auspices of Department of Health and Human Services Task Force on Alzheimer’s Disease. Neurology. 1984;34:939-.

[36] Dubois B, Feldman HH, Jacova C, Cummings JL, DeKosky ST, Barberger-Gateau P, et al. Revising the definition of Alzheimer’s disease: a new lexicon. The Lancet Neurology. 2010;9:1118–27.

[37] Guo QH, Lu CZ, Hong Z. Auditory verbal memory test in Chinese elderly. Chinese Mental Health Journal. 2001;15:13–5.

[38] Guo QH, Hong Z, Lv CZ, Zhou Y, Lu JC, Ding D. Application of Stroop color-word test on Chinese elderly patients with mild cognitive impairment and mild Alzheimer’s dementia. Chinese Journal of Neuromedicine. 2005;4:701–4.

[39] Reitan R. The validity of the trail making test as an indicator of organic brain damage. Perceptual and Motor skills. 1958;8:271–176.

[40] Rouleau I, Salmon DP, Butters N, Kennedy C, McGuire K. Quantitative and qualitative analyses of clock drawings in Alzheimer’s and Huntington’s disease. Brain and cognition. 1992;18:70–87.

[41] Sheridan LK, Fitzgerald HE, Adams KM, Nigg JT, Martel MM, Puttler LI, et al. Normative Symbol Digit Modalities Test performance in a community-based sample. Archives of Clinical Neuropsychology. 2006;21:23–8.

[42] Guo QH, Hong Z, Shi WX. Boston Naming Test in Chinese elderly, patient with mild cognitive impairment and Alzheimer’s dementia. Chinese Mental Health Journal. 1991;20:81.

[43] Fischl B. FreeSurfer. Neuroimage. 2012;62:774–81.

[44] Esteban O, Markiewicz CJ, Blair RW, Moodie CA, Isik AI, Erramuzpe A, et al. fMRIPrep: a robust preprocessing pipeline for functional MRI. Nature methods. 2019;16:111–6.

[45] Lopresti BJ, Klunk WE, Mathis CA, Hoge JA, Ziolko SK, Lu X, et al. Simplified quantification of Pittsburgh Compound B amyloid imaging PET studies: a comparative analysis. Journal of Nuclear Medicine. 2005;46:1959–72.

[46] Wong K-P, Wardak M, Shao W, Dahlbom M, Kepe V, Liu J, et al. Quantitative analysis of [18 F] FDDNP PET using subcortical white matter as reference region. European journal of nuclear medicine and molecular imaging. 2010;37:575–88.

[47] Villemagne VL, Rowe CC. Amyloid imaging. International psychogeriatrics. 2011;23:S41–S9.

[48] Huang K-L, Lin K-J, Hsiao I-T, Kuo H-C, Hsu W-C, Chuang W-L, et al. Regional amyloid deposition in amnestic mild cognitive impairment and Alzheimer’s disease evaluated by [18F] AV-45 positron emission tomography in Chinese population. PloS one. 2013;8:e58974.

[49] Sperling RA, Johnson KA, Doraiswamy PM, Reiman EM, Fleisher AS, Sabbagh MN, et al. Amyloid deposition detected with florbetapir F 18 (18F-AV-45) is related to lower episodic memory performance in clinically normal older individuals. Neurobiology of aging. 2013;34:822–31.

[50] Shokouhi S, Mckay JW, Baker SL, Kang H, Brill AB, Gwirtsman HE, et al. Reference tissue normalization in longitudinal 18 F-florbetapir positron emission tomography of late mild cognitive impairment. Alzheimer’s research & therapy. 2016;8:1–12.

[51] Thal DR, Ru□b U, Orantes M, Braak H. Phases of Aβ-deposition in the human brain and its relevance for the development of AD. Neurology. 2002;58:1791–800.

[52] Chen K, Roontiva A, Thiyyagura P, Lee W, Liu X, Ayutyanont N, et al. Improved power for characterizing longitudinal amyloid-β PET changes and evaluating amyloid-modifying treatments with a cerebral white matter reference region. Journal of Nuclear Medicine. 2015;56:560–6.

[53] Qi X, Arfanakis K. Regionconnect: Rapidly extracting standardized brain connectivity information in voxel-wise neuroimaging studies. Neuroimage. 2021;225:117462.

[54] Schaefer A, Kong R, Gordon EM, Laumann TO, Zuo X-N, Holmes AJ, et al. Local-global parcellation of the human cerebral cortex from intrinsic functional connectivity MRI. Cerebral cortex. 2018;28:3095–114.

[55] M Tucker A, Stern Y. Cognitive reserve in aging. Current Alzheimer Research. 2011;8:354–60.

[56] Whalley LJ, Deary IJ, Appleton CL, Starr JM. Cognitive reserve and the neurobiology of cognitive aging. Ageing research reviews. 2004;3:369–82.

[57] Raz N, Ghisletta P, Rodrigue KM, Kennedy KM, Lindenberger U. Trajectories of brain aging in middle-aged and older adults: regional and individual differences. Neuroimage. 2010;51:501–11.

[58] Tucker-Drob EM. Cognitive aging and dementia: a life-span perspective. Annual review of developmental psychology. 2019;1:177–96.

[59] Tucker-Drob EM, Brandmaier AM, Lindenberger U. Coupled cognitive changes in adulthood: A meta-analysis. Psychological bulletin. 2019;145:273.

[60] Baumgart M, Snyder HM, Carrillo MC, Fazio S, Kim H, Johns H. Summary of the evidence on modifiable risk factors for cognitive decline and dementia: a population-based perspective. Alzheimer’s & Dementia. 2015;11:718–26.

[61] Steffener J, Barulli D, Habeck C, O’Shea D, Razlighi Q, Stern Y. The role of education and verbal abilities in altering the effect of age-related gray matter differences on cognition. PLoS One. 2014;9:e91196.

[62] Gazes Y, Bowman FD, Razlighi QR, O’Shea D, Stern Y, Habeck C. White matter tract covariance patterns predict age-declining cognitive abilities. NeuroImage. 2016;125:53–60.

[63] Wang D, Li X, Dang M, Zhao S, Sang F, Zhang Z. Frontotemporal structure preservation underlies the protective effect of lifetime intellectual cognitive reserve on cognition in the elderly. Alzheimers Res Ther. 2024;16:255.

[64] Goldberg E, Bilder RM. The frontal lobes and hierarchical organization of cognitive control. The frontal lobes revisited: Psychology Press; 2019. p. 159–87.

[65] Stuss DT, Benson DF. The frontal lobes and control of cognition and memory. The frontal lobes revisited: Psychology Press; 2019. p. 141–58.

[66] Taren AA, Venkatraman V, Huettel SA. A parallel functional topography between medial and lateral prefrontal cortex: evidence and implications for cognitive control. Journal of Neuroscience. 2011;31:5026–31.

[67] Kozlovskiy SA, Nikonova EY, Pyasik MM, Velichkovsky BM. The cingulate cortex and human memory processes. Psychology in Russia. 2012;5:231.

[68] Stevens FL, Hurley RA, Taber KH. Anterior cingulate cortex: unique role in cognition and emotion. The Journal of neuropsychiatry and clinical neurosciences. 2011;23:121–5.

[69] Dadario NB, Sughrue ME. The functional role of the precuneus. Brain. 2023;146:3598–607.

[70] Utevsky AV, Smith DV, Huettel SA. Precuneus is a functional core of the default-mode network. Journal of Neuroscience. 2014;34:932–40.

[71] Margulies DS, Ghosh SS, Goulas A, Falkiewicz M, Huntenburg JM, Langs G, et al. Situating the default-mode network along a principal gradient of macroscale cortical organization. Proceedings of the National Academy of Sciences. 2016;113:12574–9.

[72] Fjell AM, Walhovd KB, Fennema-Notestine C, McEvoy LK, Hagler DJ, Holland D, et al. One-year brain atrophy evident in healthy aging. Journal of Neuroscience. 2009;29:15223–31.

[73] Fjell AM, McEvoy L, Holland D, Dale AM, Walhovd KB, Initiative AsDN. What is normal in normal aging? Effects of aging, amyloid and Alzheimer’s disease on the cerebral cortex and the hippocampus. Progress in neurobiology. 2014;117:20–40.

[74] Fjell AM, McEvoy L, Holland D, Dale AM, Walhovd KB, Initiative AsDN. Brain changes in older adults at very low risk for Alzheimer’s disease. Journal of Neuroscience. 2013;33:8237–42.

[75] Buckner RL. Memory and executive function in aging and AD: multiple factors that cause decline and reserve factors that compensate. Neuron. 2004;44:195–208.

[76] Buckner RL, DiNicola LM. The brain’s default network: updated anatomy, physiology and evolving insights. Nature reviews neuroscience. 2019;20:593–608.

[77] Yang Y, Chen Y, Sang F, Zhao S, Wang J, Li X, et al. Successful or pathological cognitive aging? Converging into a” frontal preservation, temporal impairment (FPTI)” hypothesis. Science bulletin. 2022;67:2285–90.

[78] Gutchess A. Plasticity of the aging brain: new directions in cognitive neuroscience. Science. 2014;346:579–82.

[79] Greenwood PM. Functional plasticity in cognitive aging: review and hypothesis. Neuropsychology. 2007;21:657.

[80] Bishop NA, Lu T, Yankner BA. Neural mechanisms of ageing and cognitive decline. Nature. 2010;464:529–35.

[81] Grady C. The cognitive neuroscience of ageing. Nature Reviews Neuroscience. 2012;13:491–505.

[82] Cabeza R, Albert M, Belleville S, Craik FIM, Duarte A, Grady CL, et al. Maintenance, reserve and compensation: the cognitive neuroscience of healthy ageing. Nat Rev Neurosci. 2018;19:701–10.

[83] Satz P, Cole MA, Hardy DJ, Rassovsky Y. Brain and cognitive reserve: mediator (s) and construct validity, a critique. Journal of clinical and experimental neuropsychology. 2011;33:121–30.

[84] Kobayashi T, Zhang H, Tang WW, Irie N, Withey S, Klisch D, et al. Principles of early human development and germ cell program from conserved model systems. Nature. 2017;546:416–20.

[85] Opdebeeck C, Martyr A, Clare L. Cognitive reserve and cognitive function in healthy older people: a meta-analysis. Aging, Neuropsychology, and Cognition. 2016;23:40–60.

[86] Hendrie HC, Smith-Gamble V, Lane KA, Purnell C, Clark DO, Gao S. The association of early life factors and declining incidence rates of dementia in an elderly population of African Americans. The Journals of Gerontology: Series B. 2018;73:S82–S9.

[87] Manly JJ, Jones RN, Langa KM, Ryan LH, Levine DA, McCammon R, et al. Estimating the prevalence of dementia and mild cognitive impairment in the US: the 2016 health and retirement study harmonized cognitive assessment protocol project. JAMA neurology. 2022;79:1242–9.

[88] Sando SB, Melquist S, Cannon A, Hutton M, Sletvold O, Saltvedt I, et al. RiskLreducing effect of education in Alzheimer’s disease. International Journal of Geriatric Psychiatry: A journal of the psychiatry of late life and allied sciences. 2008;23:1156–62.

[89] Groot C, van Loenhoud AC, Barkhof F, van Berckel BN, Koene T, Teunissen CC, et al. Differential effects of cognitive reserve and brain reserve on cognition in Alzheimer disease. Neurology. 2018;90:e149–e56.

[90] Lovden M, Fratiglioni L, Glymour MM, Lindenberger U, Tucker-Drob EM. Education and Cognitive Functioning Across the Life Span. Psychol Sci Public Interest. 2020;21:6–41.

[91] Nyberg L, Magnussen F, Lundquist A, Baare W, Bartres-Faz D, Bertram L, et al. Educational attainment does not influence brain aging. Proc Natl Acad Sci U S A. 2021;118.

[92] Averbeck B, O’Doherty JP. Reinforcement-learning in fronto-striatal circuits. Neuropsychopharmacology. 2022;47:147–62.

[93] Kleerekooper I, van Rooij SJ, van den Wildenberg WP, de Leeuw M, Kahn RS, Vink M. The effect of aging on fronto-striatal reactive and proactive inhibitory control. NeuroImage. 2016;132:51–8.

[94] Lindenberger U, Lövdén M. Brain plasticity in human lifespan development: the exploration–selection–refinement model. Annual Review of Developmental Psychology. 2019;1:197–222.

