## Supplemental figures for "Quantifying Cognitive Reserve Through Structural-Functional Interactions: Neuroadaptive Biomarkers in Aging and Neurodegenerative Pathologies"

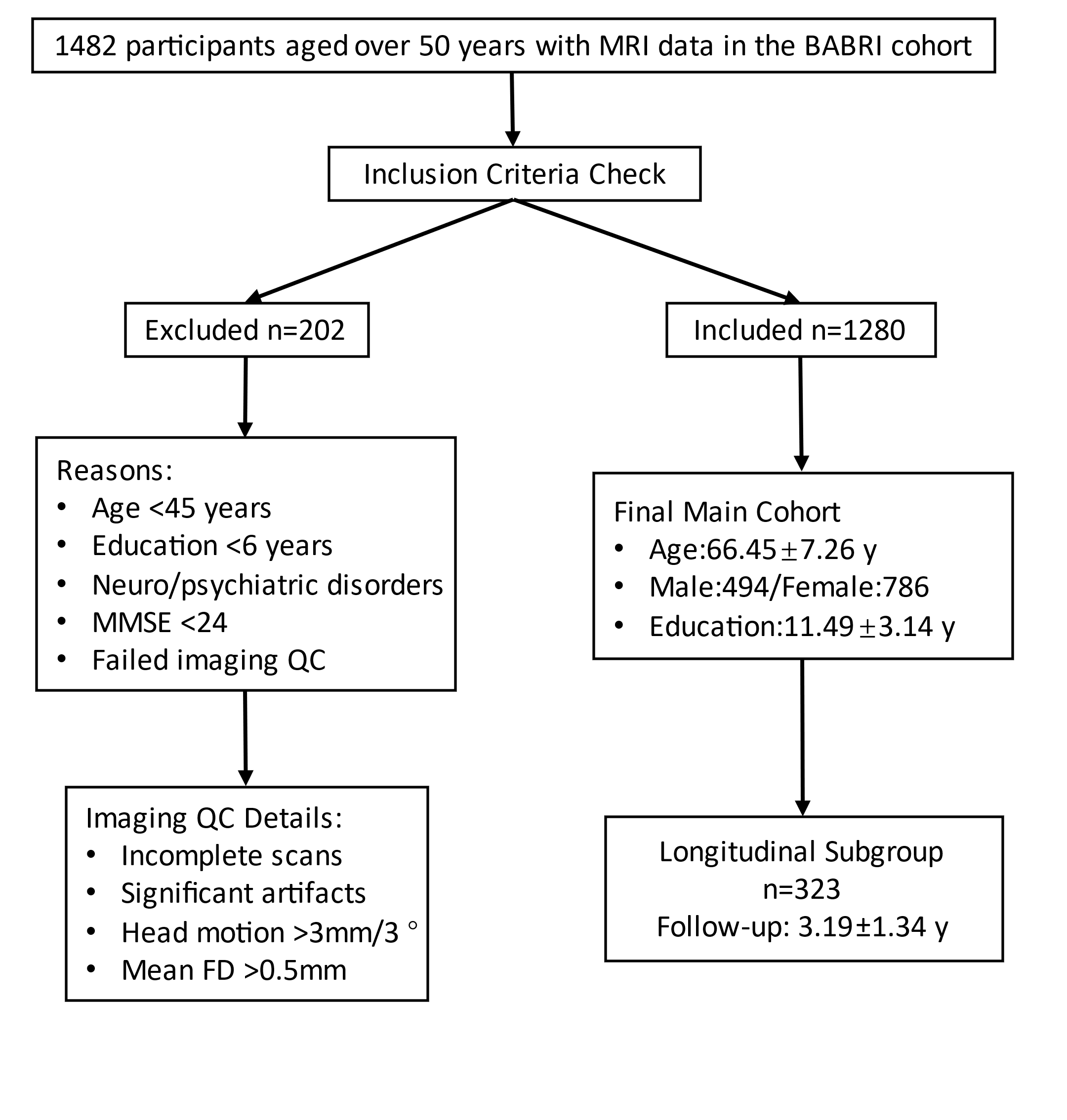

**Supplementary Figure 1. Participant Screening Flowchart of the BABRI Cohort.**
The flowchart details the inclusion/exclusion process for the community-based aging cohort. From an initial pool of 1,482 participants (aged 50–90 years), 202 were excluded due to failure to meet clinical criteria or poor-quality neuroimaging data (incomplete scans, motion >3 mm/3°, or mean FD >0.5 mm). The final analytical sample comprised 1,280 older adults (age: 66.45 ±7.26 years; 61.4% female; education: 11.49 ±3.14 years). A longitudinal subgroup (N = 323) underwent repeated cognitive assessments over 3.19 ±1.34 years.

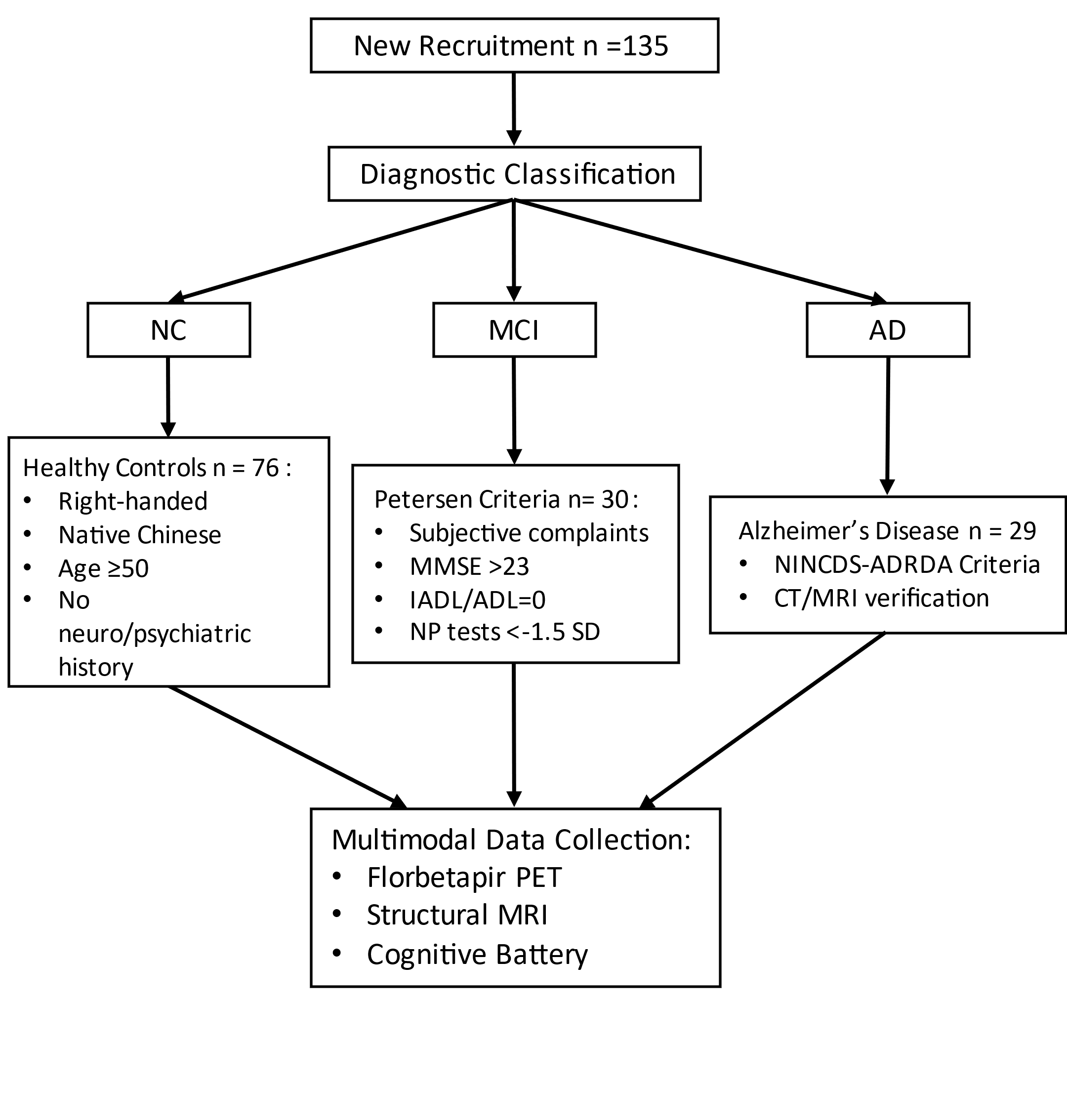

**Supplementary Figure 2. Participant Screening and Diagnostic Workflow for the Disease Cohort.** This flowchart outlines the recruitment and diagnostic pipeline for the disease-related cognitive reserve cohort (N = 135), independent of the main aging cohort. Participants included 76 cognitively normal (CN), 30 mild cognitive impairment (MCI), and 29 Alzheimer’s disease (AD) cases. **Inclusion criteria**: right-handedness, native Chinese speakers, age ≥50 years, and absence of neurological/psychiatric disorders. **Diagnostic protocols**: **MCI**: Petersen criteria (subjective complaints, MMSE >23, intact daily function [IADL/ADL = 0], neuropsychological scores <1.5 SD below age/education-adjusted norms) (Petersen, 2010). **AD**: NINCDS-ADRDA criteria with CT/MRI confirmation. All participants completed florbetapir PET, structural MRI, and cognitive assessments within one month.
